# Proteome-wide model for human disease genetics

**DOI:** 10.1101/2023.11.27.23299062

**Authors:** Rose Orenbuch, Courtney A. Shearer, Aaron W. Kollasch, Hansen D. Spinner, Thomas A. Hopf, Lood van Niekerk, Dinko Franceschi, Mafalda Dias, Jonathan Frazer, Debora S. Marks

## Abstract

Identifying variants driving disease accelerates both genetic diagnosis and therapeutic development, but missense variants still present a bottleneck as their effects are less straightforward than truncations or nonsense mutations. While computational prediction methods are sufficiently accurate to be of clinical value for variants in *known* disease genes, they do not generalize well to other genes as the scores are not calibrated across the proteome^1–6^. To address this, we developed a deep generative model, popEVE, that combines evolutionary information with population sequence data^7^ and achieves state-of-the-art performance on a suite of proteome-wide prediction tasks, without overestimating the prevalence of deleterious variants in the population. popEVE identifies 442 genes in a developmental disorder cohort^8^, including evidence of 123 novel candidates, many without the need for cohort-wide enrichment. Candidate genes are functionally similar to known developmental disorder genes and case variants tend to fall in functionally important regions of these genes. Finally, we show that these findings can be reproduced from analysis of the patient exomes alone, demonstrating that popEVE provides a new avenue for genetic analysis in situations where traditional methods fail, including genetic diagnosis of rare-as-one diseases, even in the absence of parent sequencing.

## Introduction

Even if every human were sequenced and their phenotypes recorded, the space of disease-causing genetic variation may be too large to be studied by population variation or disease relevant experimental assays alone ^9,10^. Patients with unique combinations of symptoms and genotypes would still go without a genetic diagnosis^9,10^. The biodiversity of life on Earth provides a deeper view of genetic variation across billions of years of evolution, presenting a unique opportunity to uncover complex genetic patterns preserved to maintain fitness. Thus, models that can distill such information can accelerate our ability to leverage genetics for diagnosis, preventive care, and treatment.

In the context of severe genetic disorders, the task is to identify the causal variant amongst the millions of mutations in a single individual. One powerful approach is the sequencing of trios – patient and their parents – which provides a way to narrow down the candidate variants to those arising *de novo* when the parents are thought to be unaffected or inherited variants from an affected parent^11,12^. However, despite impressive analysis of large rare disease cohorts^8,12–16^, genetic diagnostic yield is relatively low; in some cases only 25% of probands receive a genetic diagnosis^16^. There is a need for alternative strategies to identify candidate causal variants directly from a patient’s sequencing data, without relying on frequency of observations in large cohorts. In this work, we present how probabilistic modelling of diverse sequencing efforts, in both human and across diverse species, can provide an answer with the potential to enable clinical interpretation of never-before-seen variants.

Recent work using deep unsupervised models trained only on evolutionary sequences have shown strong promise for clinical variant effect prediction^1–6^ and have demonstrated comparable accuracy to experimental approaches^1^. Since these models do not depend on functional or clinical labeling, they can generalize to variants in genes without previous annotation. However, although these models often perform well in terms of separating *Benign* from *Pathogenic* clinical labels in known disease genes, they are not calibrated well across the entire human proteome – i.e. they are not designed for comparing how deleterious a variant is in one gene versus a variant in another. Consequently, previous methods excel at identifying variants that disrupt the function of the resulting protein but do not necessarily predict if it is detrimental at the organismal level^17^.

Variant severity lies on a spectrum: disruption of function in one protein could have modest effects late in life, while the disruption of another protein can be lethal in childhood, for instance. Both can be considered “pathogenic” and correctly identified as such by a model, but when attempting to find the genetic cause of a severe disorder, it is imperative to be able to distinguish between these two scenarios. Current state-of-the-art variant effect prediction models have not been developed with this spectrum of severity in mind.

To overcome this, we developed popEVE, a model that places variants on a proteome-wide scale of deleteriousness, enabling us to predict if a variant seen in one gene is more detrimental to human health than a variant seen in another. popEVE leverages deep evolutionary data to achieve missense resolution variant effect prediction and shallow variation across the UK Biobank^7^ or GnomAD v2^18^ population to transform the score to reflect human-specific constraint. Analyzing a meta-cohort^8^ of patients with severe developmental disorders (SDD), we find evidence for at least 123 novel genetic disorders from their de novo missense variants -- 44% more than previously identified in the same cohort, and yet significantly similar in function to known developmental disease genes. For cases with whole exome sequencing, we find that in 98% of cases, the previously identified de novo variant is predicted to be the most deleterious variant across the whole exome. Thus, popEVE provides valuable information for genetic diagnosis, even in the absence of trio sequencing, increasing the scope of genetic analysis.

## Results

### A unified model of population and evolutionary sequences

For a computational model to be broadly useful in human genetics, the model scores should be continuous, have residue resolution and have the same quantitative meaning across different proteins. Previous state-of-art computational methods have excelled in various tests of accuracy; for instance, correct classification of pathogenic and benign labels from curated clinical databases and reasonable correlations with high throughput experiments on specific proteins^1,19–23^. However, these benchmarks can result in overestimated accuracy in and generalizability to real-world scenarios in which thousands of missense variants, including hundreds of rare variants, must be ranked across a single person’s genome. This drawback has resulted in the understandable caution of the clinical use of computational methods, not least from the observation of an over-prediction of deleterious variants^19,23–26^.

### Converting gene-level scores to proteome-wide scores

By contrast popEVE is designed to provide a human-specific, continuous measure of variant deleteriousness that enables realistic comparison across different proteins (Fig. 1a, Supp. Fig. 1, Methods). To achieve a score that reflects constraints within humans and yet has the resolution to distinguish the relative impact of individual variants, we reasoned that a model would need to not only learn from deep evolutionary variation but also from shallow variation observed in the human population. Whilst deep evolutionary sequence variation from across life can inform us about what is allowed for a protein to function, the models trained only in that way cannot necessarily learn the relative importance of one protein versus another. We therefore train the model to predict the presence of a variant in the UK Biobank, dependent on the underlying evolutionary model score using a latent Gaussian process prior (Methods), similar in spirit to gene-level and region-level estimates of missense constraint^27–30^. The model therefore trains on the universe of sequences across the whole of evolution together with summary statistics of human variation from human population data. For the deep evolutionary sequence analysis, we combine a state of the art alignment-based models, EVE^1^, and a large language model, ESM1v^31^. Although the two models have comparable performance on clinical label and DMS benchmarks, variant scores are not well correlated (Notin et al^32^ and Supp. Fig. 2, 3 and 4) – indicating useful orthogonal evidence of variant fitness.

**Figure 1.**
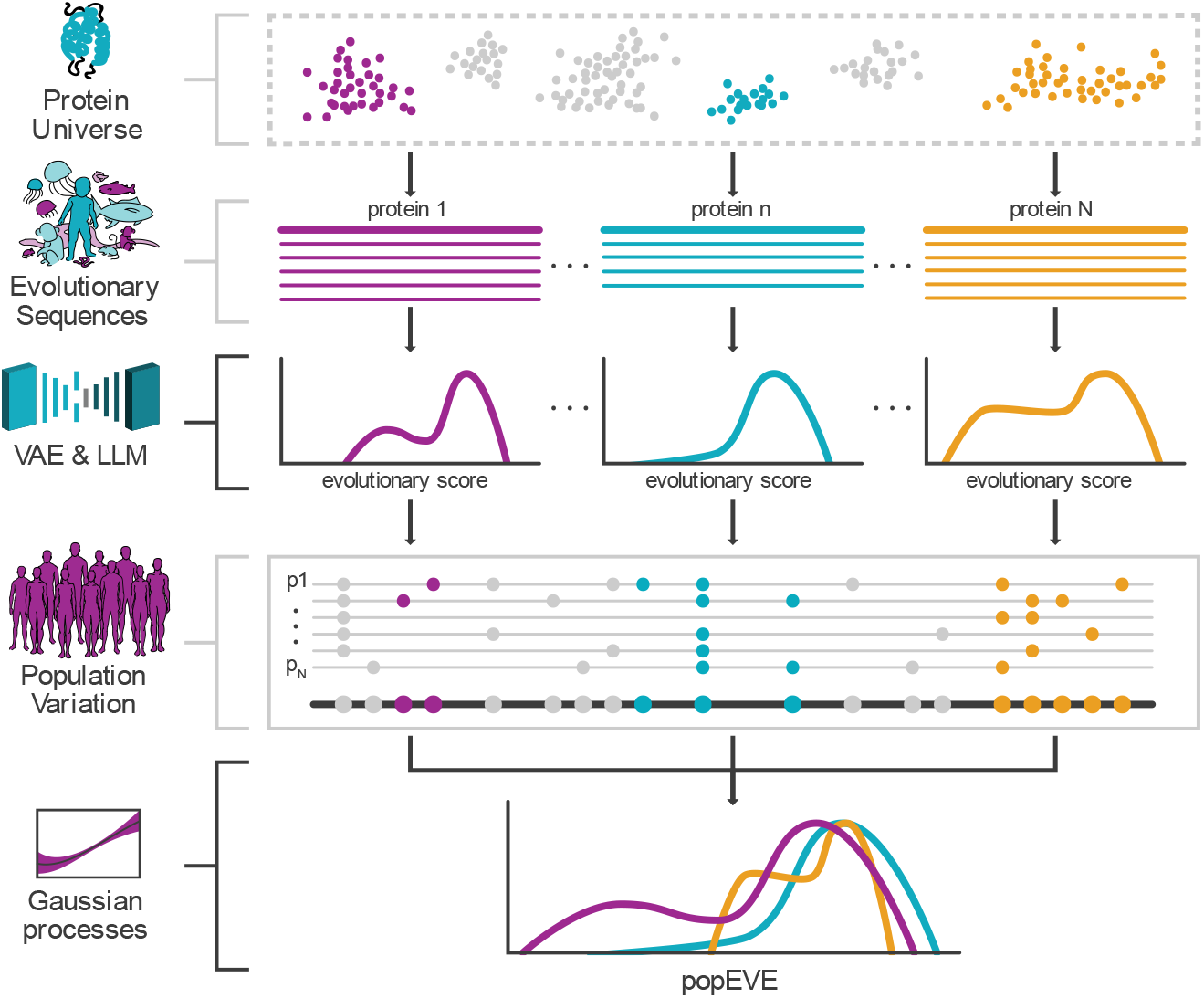
popEVE combines deep evolution and human variation. popEVE combines variation from across evolutionary sequences, modeled with EVE and ESM1v, with variation within the human population (UK biobank^1^), using a gaussian process to learn the relationship between evolutionary scores and missense constraint.

### popEVE shows limited to no population bias

We do not expect our population-adjustment framework to introduce population structure bias into our variant scores because we use a coarse measure of missense variant constraint for each gene rather than directly using allele frequencies. As such, the presence of a rare variant in a single person in the training population is treated the same as the presence of a common variant in the vast majority of people. We find that popEVE score distributions of rare variants (MAF < 0.01) are similar across various ancestries in GnomAD v2^18^ (Supp. Fig. 5). An independent analysis of ancestry bias in variant scoring methods found that popEVE shows minimal bias towards European ancestries in line with population-free methods. By contrast, state-of-the-art competitors, including AlphaMissense^20^, BayesDel^33^ and REVEL^34^, show significant bias towards these populations^35^.

### popEVE captures variant severity and pathogenicity

To be an effective tool in the human population, a variant scoring method should have missense resolution scores that capture some notion of variant fitness, deleterious, severity, penetrance, and/or pathogenicity and can be compared across the genome. While the primary advance of popEVE relates to the latter, our framework provides a novel method for ensembling variant scoring methods, modestly boosting popEVE’s performance on within-gene benchmarks related to variant pathogenicity and fitness over its constituent parts. popEVE performs competitively with other state-of-the-art models on separating Benign and Pathogenic ClinVar labels and correlation with high-throughput experimental assays (Supp. Fig. 2 & 3, Supp. Table 1). Importantly, we do not see an inflated performance estimate on ClinVar labeling tasks unlike models that directly incorporate allele frequencies; this is expected as the method alters rankings across distinct genes while leaving internal variant rankings essentially unaltered. An additional benefit of this is that popEVE can be safely incorporated into variant annotation pipelines^36^ that treat population data as an independent source of evidence without double-dipping.

Gold standard approaches to assessing variant scoring have placed great emphasis on segregating variants as either benign or pathogenic – i.e whether a variant can cause or increase risk of any disease or disorder. While this binary approach to variant interpretation has some benefit for clinical decision making, it obscures the fact that some variants will lead to more severe disease phenotypes than others. Here, we investigate popEVE’s ability to distinguish between deleterious variants that are likely to cause severe disease from those that do not.

In general, deleterious popEVE scores are enriched in haploinsufficient genes as compared to loss of function tolerant ones, and, to a lesser extent, genes tied to phenotypes with dominant inheritance patterns versus recessive ones – expected due to adjusting based on variant constraint (Supp. Fig. 6). As expected, gene-level metrics calculated from popEVE scores correlate more with missense-based constraint metrics than truncation- and loss-of-function-based metrics (Supp. Fig. 7, Supp. Table 2). Additionally, ClinVar pathogenic variants associated with diseases that have early onset or known to cause death in childhood are more deleterious according to popEVE than adult-onset or death in adulthood (Fig. 1a). The ability to distinguish childhood death/onset from adult death/onset indicates that popEVE scores contain some information on variant severity and deleteriousness in disease. In contrast, AlphaMissense, a top variant scoring method fine-tuned for predicting variant “pathogenicity”, does not distinguish these sets of variants from each other but correctly classifies most of these variants as possibly pathogenic (Supp. Fig. 8).

### Deleterious popEVE scores are enriched in severe developmental disorder cases

To further investigate our ability to capture variant severity, we compare variant scores to unaffected controls: unaffected siblings from Autism Spectrum Disorder cohort trios^37^ and individual participants in the UK Biobank (Supp. Table 3). popEVE scores are shifted towards the deleterious end in cases in comparison to controls (Fig. 2b top). With increasingly deleterious popEVE scores, we find these de novo missense variants are increasingly enriched above what we would expect given the background mutation rate (Methods, Fig. 2c top). Subsetting SDD cases to those that can be diagnosed based on their missense de novo variants following definitions in Kaplanis et al^8^ further widens the shift between the two distributions (Fig. 2a bottom). To establish a score threshold for classifying DNMs as candidates for causing disease, we fit a two-component Gaussian mixture model to the distribution of popEVE scores across all cases and controls without using their labels (Fig. 2d, Methods). We selected a high confidence severity threshold (−5.056) where 99.99% of the variant scores are in the low fitness distribution. These 1,163 variants are over 15-fold enriched in the SDD cohort versus the expected number of popEVE-severe DNMs given the background mutation rate (Fig. 2c bottom), a five times higher fold enrichment than those reported by other state-of-art methods, e.g. PrimateAI-3D at 2-fold as reported in Gao et al 2023^21^. Furthermore, even variants we define as moderately pathogenic are five times enriched in the developmental disorder cohorts, again outperforming previous methods. Notably, none of the case variants predicted to be severe or moderate are seen in the UK Biobank or GnomAD.

**Figure 2.**
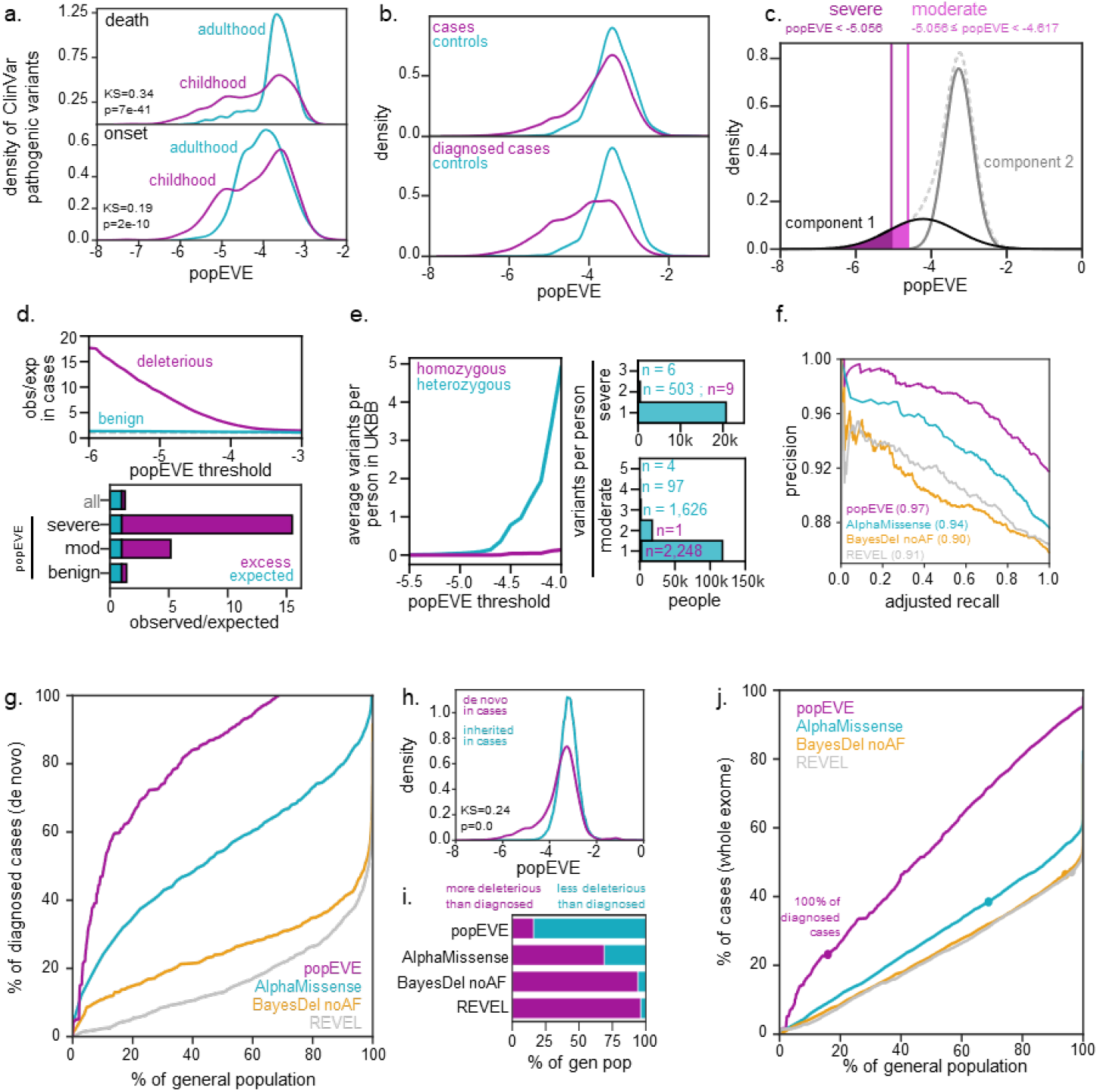
popEVE captures variant severity and pathogenicity. a. ClinVar pathogenic variants in phenotypes associated with early onset and premature death in childhood have more deleterious popEVE scores than those associated with later onset and death after maturation. There is a greater separation between the distribution of variant scores for age of death (KS=0.34, pvalue=7e-41) than onset (KS=0.19, pvalue=2e-10). Onset and death labels were acquired from OrphaNet. b. popEVE scores for de novo variants in severe developmental disorder cases (top) and diagnosed cases (bottom) are shifted towards the deleterious end compared to controls (unaffected siblings from ASD family cohorts). c. Using de novo variants from both SDD cases and controls, we define a severely and moderately deleterious score threshold by fitting a two-component gaussian mixture model and finding the 99.99% and 99% likelihood of being in the more deleterious distribution. d. With increasingly pathogenic thresholds, de novo variants in the SDD metacohort are significantly enriched (top). At our severely pathogenic threshold, popEVE pathogenic variants exhibit over 15-fold enrichment while popEVE benign variants are in line with expectation (bottom). Moderately pathogenic variants are enriched 5-fold. Expected number of variants are quantified using a background mutation rate based on the number of individuals in the metacohort. e. In the UKBB, individuals have at most one homozygous and up to three heterozygous severely deleterious variants – 96% of the 500k individuals have no severely pathogenic missense variants (left). Approximately 72% of UKBB individuals have no severely nor moderately deleterious variants and at most five moderately deleterious variants (right). f. popEVE is better at separating diagnosed developmental disorder cases from controls based on de novo mutations than other state of art variant effect predictors with an average precision of 97%. Recall is adjusted based on the expected number of these cases to have a causal de novo missense variant (See Methods). g. popEVE recalls more severe developmental disorder diagnosed cases based on their de novo variants without overpredicting pathogenicity in whole exome sequencing from relatively healthy controls from UKBB (gnomAD-trained popEVE). h. De novo missense variants in SDD cases from the DDD are enriched for pathogenic variants in comparison to their rare inherited variants (MAF<0.01) (KS=0.24, pvalue=0.0). i. To recall 100% of de novo missense diagnosed cases using their whole exome sequencing, popEVE predicts far less of the general population will have a similarly deleterious variant than any other model. j. When applied to WES from a subset of the severe developmental disorder cases, popEVE recalls more cases than other models without overpredicting pathogenicity in the general population of UK Biobank (gnomAD-trained popEVE). Additionally, popEVE recalls 100% of cases expected to have a causal de novo missense variant for only 15% of the remaining cases and 16% of the general population (circles). Other models find that over >78% of the UKBB has a variant as deleterious as these cases (inset).

### popEVE is better at distinguishing severe developmental disorder cases from controls than other variant scoring methods

To assess performance at ranking variants across the proteome, we tested our model’s ability to separate de novo mutations from missense diagnosed SDD cases from those in unaffected controls. popEVE performs better than all other state-of-the-art models at distinguishing diagnosed cases from healthy controls, improving average precision by 32% over the next best model (Fig. 2f). Notably, popEVE differentiates diagnosed cases from controls better than variant scoring methods that train directly on clinical labels that likely include diagnostic variants from these cases (Fig. 2f, Supp. Fig. 9 a-c, Supp. Table 1, Methods). Indeed, an independent analysis supports popEVE as the leading variant scoring method in identifying likely causal variants in SDD cases^17^.

### popEVE recovers severe genetic disorder cases without overpredicting severity in the general population

A model that captures severity should be able to rank variants in severe genetic disorder cases as more deleterious than those found in individuals with less severe phenotypes. As such, we compare variant scoring method’s ability to distinguish these monogenic cases thought to be caused by a single de novo missense mutation from individuals with primarily complex and adult-onset diseases in the UK Biobank (Fig. 2 g). Based on decreasingly deleterious score thresholds, popEVE recovers far more diagnosed cases without overpredicting severity in the general population. While popEVE recalls 50% of cases along with only 11% of the UK Biobank, the next best model, AlphaMissense, predicts 44% of the general population will have at least one variant as deleterious as 50% of diagnosed SDD cases. Furthermore, at this threshold, AlphaMissense predicts an average of five “pathogenic” variants per person while popEVE returns far less than one per person on average.

As a final performance assessment, we tested how well models differentiate SDD cases from controls using their whole exome sequencing (WES) – both inherited and de novo variants. The distribution of inherited variants is similar to variants seen in the UK Biobank participants while de novo variants are shifted towards the deleterious end (Fig. 2 h). For a subset of the SDD cases with WES, popEVE finds a primarily 1:1 relationship between cases and controls except for the most deleterious of thresholds (Fig. 2 i-j, Supp. Fig. 9e, Methods). Strikingly, the inflection point of this function lies at the threshold where popEVE recovers 100% of SDD cases that are expected to have a causal de novo missense variant. Other methods vastly overpredict the amount of deleterious variants in the general population compared to WES from cases and predict that nearly 100% of the UK Biobank has variants as severe as half of these cases. We find, again, that popEVE outperforms other models at recovering more severe developmental disorder cases for fewer healthy controls.

### Evidence of 123 novel candidate developmental disorder genes

Given its performance across the various benchmarks and lack of biases, popEVE appears uniquely suited for use in clinical genetics settings to identify candidate variants. We first investigated popEVE’s utility in discovering novel variants and genes in the SDD cohort, comprised of 31k trios in total (Supp. Table 3).

### Candidate variant and gene discovery

We used a two-pronged approach to discover associations: (1) thresholding the scores for more than a 99.99% likelihood of falling within the low fitness distribution described previously (2) gene collapsing, comparing variant scores seen in the cohort to what is expected given the background mutation rate and the spectrum of popEVE scores within and across proteins (p<2.71×10^−6^, Methods). This results in 442 genes including 183 that were identified by a previous study of the same cohort using DeNovoWEST^8^ (Fig. 3 a, Supp. Table 4). popEVE recalls 94% of genes previously identified in the three cohorts and over half (135) of the new gene set have been associated with developmental disorders from other cohorts according to the Developmental Disorder Gene to Phenotype (DDG2P) database ^38^. 123 of the genes are novel candidates – 119 of which were identifiable at the single-variant level (Supp. Table 5). Interestingly, we recover 31 genes using missense variants alone whereas previous work only identified the gene using patients with LoF and missense variants. Of the 50 previously known genes that we missed by variant alone but recalled with gene collapsing, many have moderately deleterious scores, highlighting the benefit of using a two-pronged approach.

**Figure 3.**
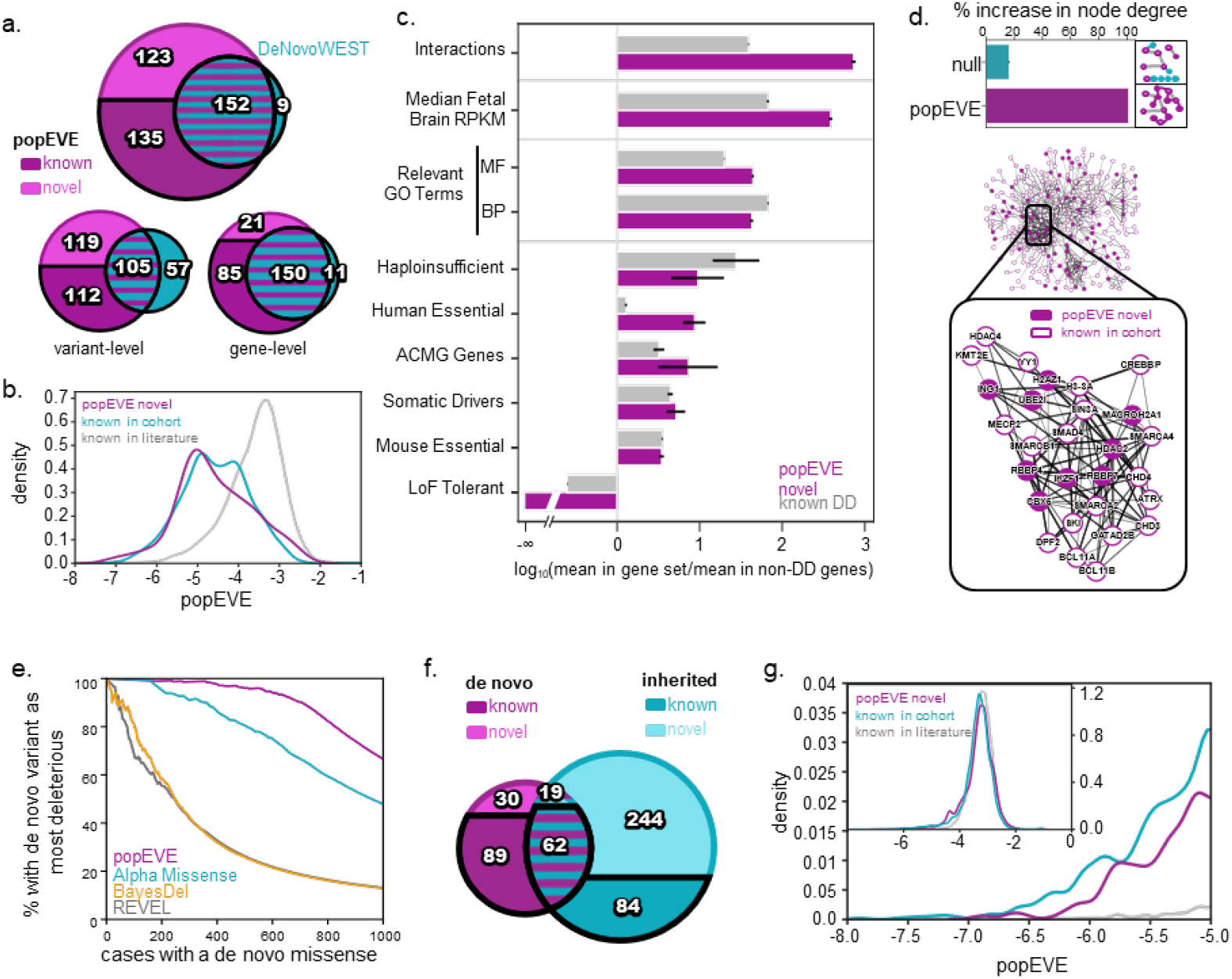
popEVE finds evidence for 123 novel candidate genes in severe developmental disorders. a. Both popEVE gene and variant-association methods achieve an 80% recall when compared to the DeNovoWEST^8^ analysis of the SDD metacohort and a 95% recall of the genes discoverable by DNW with solely missense variation. There is a greater overlap between popEVE gene-collapsing and DNW missense than the thresholding approach, due to the similarity in their approach. b. Novel candidate genes and genes previously identified in the same SDD cohort have a similar distribution of de novo case variants as compared to developmental disorder genes identified elsewhere. c. Novel and known developmental disorder genes show similar enrichment in properties known to differentiate known DD-genes from non-developmental disorder genes (95% CI from bootstrapping shown). d. Novel genes have many biochemical interactions11 with genes previously identified in the same cohort - inclusion of novel genes increases node degree 100% compared to random sets (two-sided T test, p=0) (top). The densest portion of the network (middle) includes many genes involved in chromatin modeling (bottom). e. popEVE ranks de novo missense variant in known developmental disorder genes as most deleterious compared to their inherited variants in diagnosed cases, better than any other model. f. Genes identified using de novo variants overlap with those identified using inherited variants. g. Novel candidate genes and genes previously identified in the same SDD cohort have a similar distribution of whole-exome case variants as compared to developmental disorder genes identified elsewhere, particularly at the deleterious end. (Inset shows entire distribution of scores).

Taken together, we provide evidence that variant score thresholding alone can provide accurate results and that it is beneficial to use both methods when possible.

### Functional analysis supports candidate genes

Three broad lines of evidence provide support for new candidates. Firstly, 70% of the 410 genes identified using popEVE in the SDD cohort are already known to be associated with developmental disorders (p<0.001 compared to random, Fig. 2b). Additionally, the popEVE scores distribution of variants seen in cases in the candidate genes is near identical to those in already known genes (Fig. 3b, Supp. Fig. 10).

Secondly, we found our candidate genes to be functionally similar to known DD genes across a wide range of features known to distinguish these genes from those not associated with developmental disorders (Fig. 3c, Supp. Table 6). For example, candidate genes are expressed significantly more in the developing fetal brain as compared to non-DD genes than even those already known to be associated with developmental disorders (p<0.001). Furthermore, many of these genes are involved in neurogenesis and neuronal differentiation, including PPP3R1 and PPP3CB – both are components of calcineurin, a regulator of calcium signaling and neuronal plasticity^39^. Beyond these areas, novel genes have similar enrichment for molecular function and biological processes from Gene Ontology annotations^40,41^ relevant to known developmental disorder genes, such as chromatin organization (GO:0006325) and nervous system development (GO:0007399) (Supp. Table 7). Additionally, novel candidate genes are enriched for essential genes measured both by homology to mouse experiments^42^ and by large scale CRISPR screens^43^, somatic driver genes^44^, and haploinsufficient genes^45^. Finally, novel genes are significantly depleted of genes that are tolerant to homozygous LoF or those with recessive inheritance patterns than known-DD genes. This is expected as these are candidate genes for disorders suspected to be monogenic and likely to be caused by a single de novo variant.

Thirdly, perhaps the strongest support for these candidates come from the number of direct physical interactions with the 285 genes previously identified by Kaplanis et al^8^ in the same cohort, (direct interactions 65/123 and indirect another 34). When compared to random, our novel discoveries double the density of interactions when added to the network of 285 genes (Fig. 3d, Supp. Fig. 11 & 12a, Supp. Table 8). For example, the densest portion of the network contains the NuRD chromatin complex which is crucial in embryonic development. This complex contains several genes associated with developmental disorders, including CHD3/4 and MTA. We identify five new genes in NuRD complexes including HDAC2/5, RBBP4/7, and IKZF1 and five in another chromatin associated complex, Sin3 complex (Supp. Table 8). HDAC2 itself interacts with 8 of the 285 previously identified genes and the most deleterious variant in the cohort, M31R, lies directly in the ‘foot pocket’ of the acetylase active site^46^ (Fig. 4a). The top three most deleterious DNMs in novel candidate genes, ETF1, RBBP4 and WDR5, have direct interactions to 9, 15 and 6 DD genes respectively (Supp. Table 8). Taken together, the novel genes overwhelmingly interact with genes already identified in the same cohort or elsewhere in the literature. For instance, 50/124 novel genes interact with NRTK1, a complex involved in development and survival of neurons, 16/124 interact with SMARCA4 in the SWI/SNF chromatin remodeling complex associated with neurodevelopmental disorders^47,48^, another 15 are in ion channel complexes^49^ (Supp. Table 7). Of the 24 remaining genes with no connectivity, two thirds are significantly enriched in annotations for neuronal development and differentiation (Supp. Table 7).

**Figure 4.**
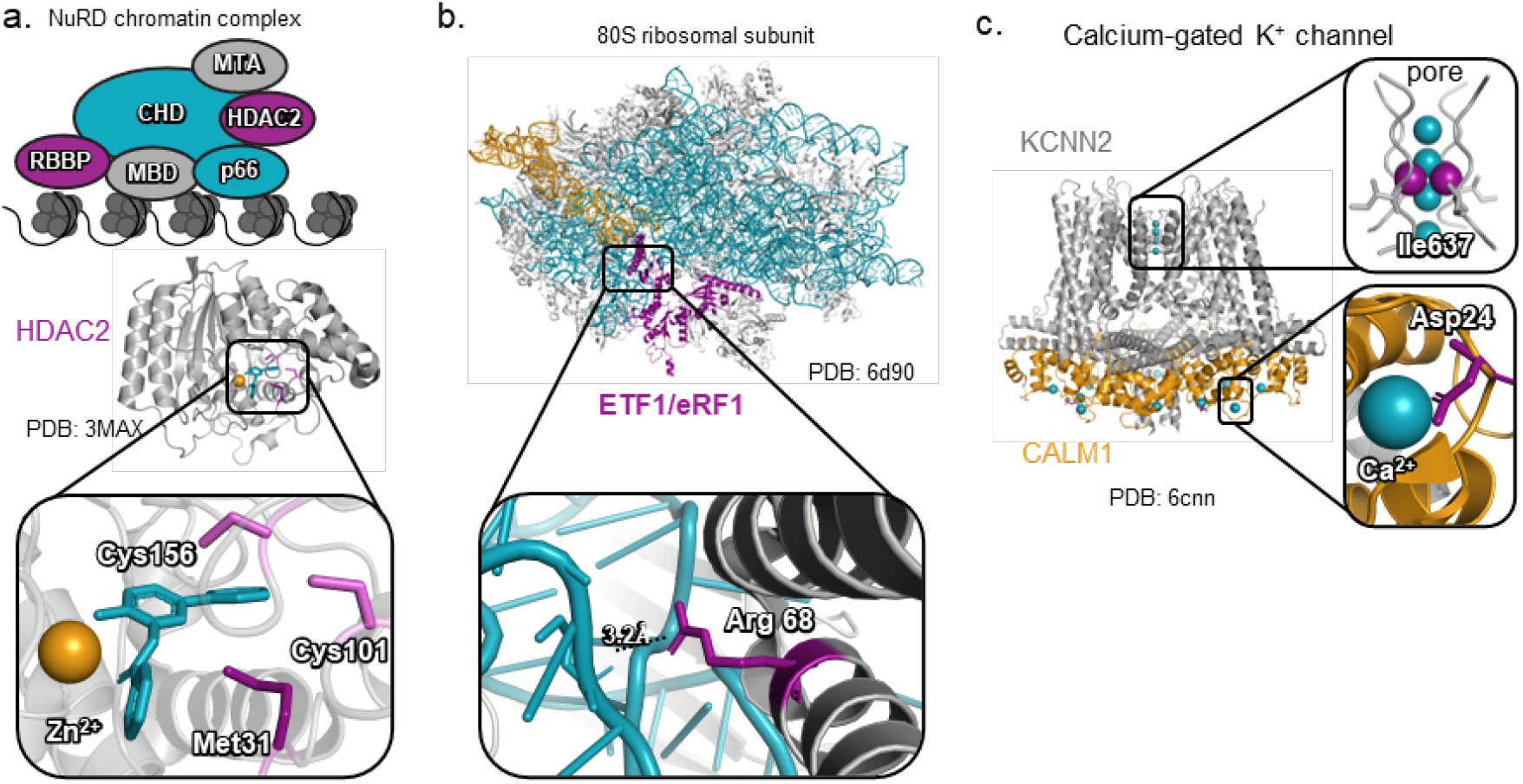
Case variants lie in functionally important regions of candidate genes a. The novel gene, HDAC2, interacts with many genes known in severe developmental disorders including those in the NuRD complex where the variant Met31Arg is proximal to the foot-pocket of the active site (PDB:3max12). b. The top scoring variants Arg68Leu and Arg192Cys in ETF1/ erF1(a translation termination factor) are contacting the anticodon site and the peptidyl transferase site in the ribosomal RNA, ternary complex (PDB:6d9013) c. novel discoveries KCNN2 and CALM1 both contain highs scoring variants in functional sites - Ile637Phe in the highly conserved T(V/I)GYG pore motif and Asp24Tyr in CALM1 which chelates the Ca2+ in the wild type (homologous complex structure PDB: 6cnn14) see Table 1.

**Table 1.**
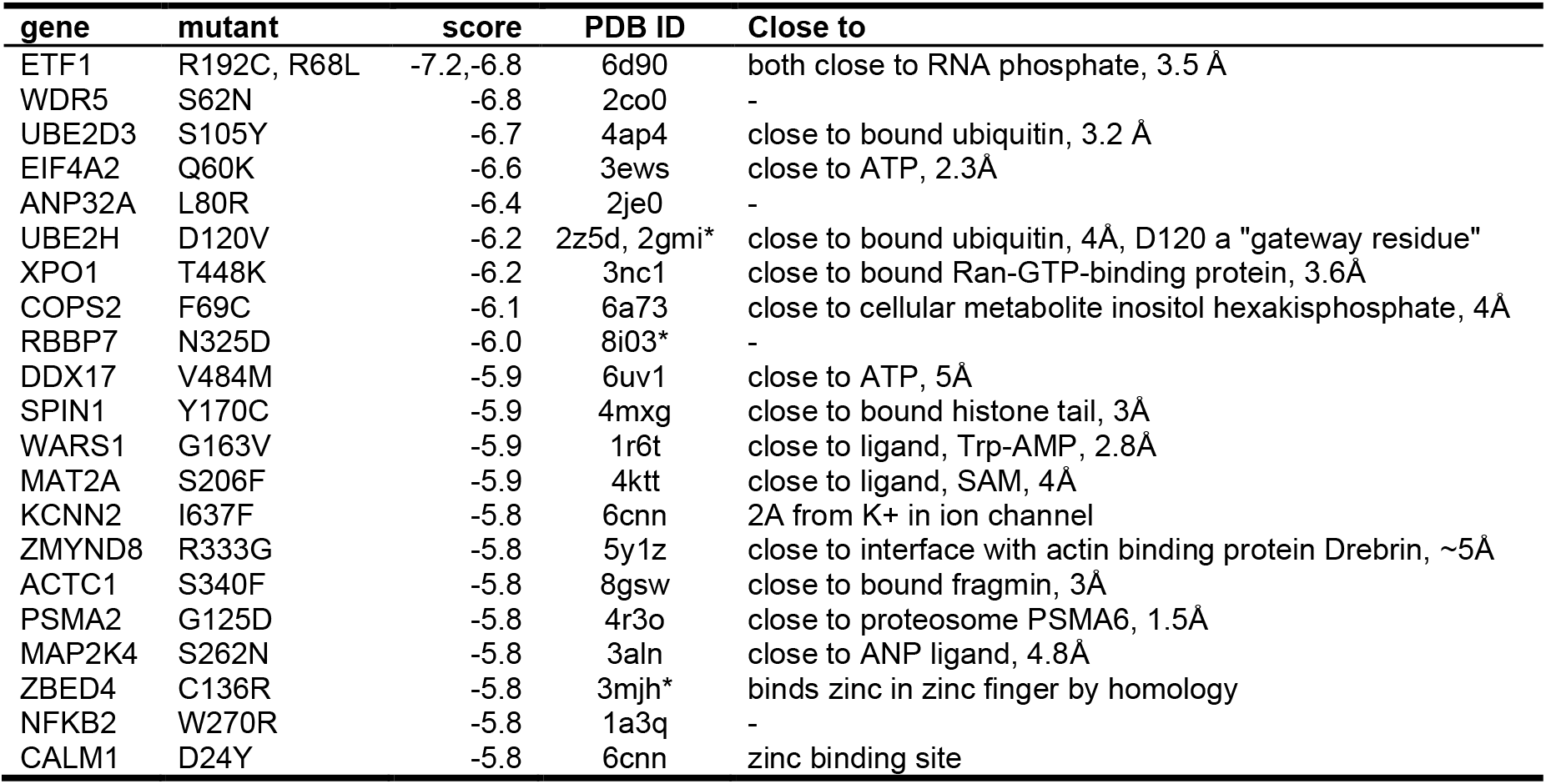
Top 20 most pathogenic novel discoveries.

These three lines of evidence taken with the low potential of false positives (see section on severity inflation in the general population) support these novel genes’ candidacy for their involvement in severe developmental disorders.

### Case variants lie in functionally important regions of candidate genes

Candidate variants tend to fall in functionally important regions of proteins. We assembled a database of 3D structures from the PDB^50^ and assessed the variant’s distance to interacting partners, e.g. metal, ligand, cofactor, protein (Methods). In the 100 candidate genes with a structure, we find that 91% and 72% of variants are within 8Å and 5Å respectively of an interaction partner (Supp. Table 9). Strikingly, these variants are 92% closer to the ligand than any other position in their respective protein according to a null model (Methods). For example, the top two most deleterious candidate variants, R192 and R68, in ETF1 (eRF1) are close (3.2Å) to the phosphate backbone of RNA in the eRF1-eRF3-GTP ternary complex that mediates translation termination (Fig. 4b, Table 1), 6d90^51^; In the calcium gated potassium ion channel, KCNN2 (Fig. 4c, modeled on KCNN1, PDB: 6cnn^52^, Table 1). Ile637 is part of the T(V/I)GYG pore motif that is essential for ion transport. Another top variant, D24Y in CALM1 (PDB: 6cnn^52^) has a mutation that is 2.4 angstroms from the activating Ca^2+^.

### Pinpointing likely-causal de novo variants without parental genomes

Finally, we asked whether our results can be obtained directly from the child genome alone without parental genetic information. To test our model’s ability to identify causal mutations without de novo labels, we investigated rare variants (MAF < 0.01), both inherited and de novo, in the subset of the SDD cohort with available whole exome sequencing, almost 10k individuals from the Deciphering Developmental Disorders study^14^ (DDD). For a variant scoring method to be useable in the clinical setting, the model must be able to rank variants within individual cases such that the most likely causal variant rises to the top. For 2.7k of these cases, this is expected to be a de novo missense variant. In fact, 98% of the 513 individuals with a popEVE severe de novo missense has this variant ranked as their most deleterious. Additionally, if we take the most deleterious variant per person, we still recover 95% of the genes identified by variant thresholding on the de novo variants alone, further demonstrating popEVE’s power to identify the likely most deleterious mutation. In comparison to other models, popEVE is better at ranking predicted-deleterious de novo variants as more deleterious than all rare inherited missense variants in individual cases (Fig. 3e, Supp. Fig. 9f, Methods). In other words, if an SDD patient has a likely-causal de novo missense variant, popEVE will rank this variant as the most deleterious more of the time than any other model. Not only does this test indicate a model’s utility in clinical genetics, but also evaluates its performance across the entire proteome.

### New candidates that may be inherited

Despite this shift, we nevertheless find 244 inherited variants predicted to be severely deleterious in 402 genes, only one of which is seen in the UKBB population. Of the genes with severe inherited variants, 38% of genes are already known to be associated with developmental disorder and 29 are amongst the novel genes discovered in the full SDD cohort. Genes with predicted severe inherited variants lie in similar functional pathways, increase node-connectivity to a similar degree and harbor candidate variants that are similarly close to interacting partners as genes with predicted severe de novo variants (Supp. Fig. 10 & 12b, Supp. Table 9 & 10). Additionally, case variants in these candidate genes have a similar popEVE distribution to those in genes already identified in the larger cohort (Fig. 3g). While de novo missense variants are the likely culprit for a number of these cases, inherited variants may also contribute to severe developmental disorders. Additionally, in the original family-trio analysis of this cohort, 84% of flagged variants were inherited, with the vast majority of these being missense variants^13^.

## Discussion

Patient sequencing has become standard for many diseases in several countries, with growing accessibility worldwide. Hence, there is an urgent need for variant interpretation strategies that are broadly applicable and can provide guidance even in cases where just one individual is suspected to have the disease. When studies manage to enroll enough individuals with the same rare disease, standard methods of genetic burden and enrichment become viable for discovering novel gene-disease associations. However, there remains a long tail of diseases which are so rare that such methods may never be applicable. In this work, we developed a model to aid in the genetic diagnosis of patients residing in this tail.

In recent years, there has been a surge of models capable of predicting whether variants are benign or pathogenic. However, in this area, consideration for the heterogeneity of severity and penetrance of disease-causing variants has been largely absent. Here, we explored the possibility that, in some situations, it may be beneficial to consider variants as lying on a spectrum of pathogenicity. To capture this spectrum, a model must be capable of ranking variants both within and across distinct genes, i.e., a model of the whole proteome. While several models provide proteome-scale predictions, to our knowledge, popEVE is the first to be built specifically to calibrate scores to be comparable across genes, and hence, may be regarded as the first, albeit simple, model of the human proteome.

To advance whole proteome modeling, there is a long road of necessary future developments. The next, perhaps most obvious, step is to account for protein-protein interactions, analogous to the development of protein-level models. Where early models considered each position in the sequence as statistically independent (e.g. column conservation models using MSAs), only later did models account for epistatic and higher-order interactions. Another clear limitation of this model as a representation of the whole proteome is the inability to assess loss of function variants, such as nonsense or truncation mutations, and, thus, are unable to compare their severity to missense variation. To the best of our knowledge, no unified model of loss of function and missense variants with sufficient predictive power currently exists. We note, however, that due to the modular nature of the model, it would be straightforward to incorporate such a model, should one become available in the future. In other words, the human proteome-calibration underlying popEVE is independent of the form of genetic variation and can easily be expanded.

Despite the simplicity of popEVE, it presents multiple opportunities for diagnosis and studying the genetic underpinnings of disease more broadly. Combining multiple lines of evidence, we find evidence for novel developmental disorder gene candidates that would not be discoverable by enrichment-based methods alone in this size cohort. 104 of our candidates have flagged variants in only one or two patients. Through complementary functional, structural and network analysis, we find many of these genes are intimately related to genes whose role in developmental disorders is already established and that the identified variants reside in functionally critical locations, providing further evidence that variants in these genes can indeed give rise to genetic disorders. More broadly, the model predicts that a large number of genes are capable of giving rise to severe phenotypes, implying that there are still many genetic disorders yet to be identified or even seen. A similar conclusion is reached in Kaplanis et al^8^ but via a distinct analysis. Here we clarify this forecast by predicting which genes and variants are most likely to be involved.

Finally, we must note the detrimental impact of building large-scale proteome or genome models; we are reaching a point where energy and computational consumption of developing and training models is costly both financially and environmentally^53^. In this work, we sought to use a modular approach, enabling us to repurpose previous models, as well as easily update components of the model with future developments at a minimal computational cost. Deep learning strategies with these properties are currently scarce, and we urgently need more techniques that lend themselves to reducing computational costs or have components that can be readily reused or recycled.

## Data Availability

All data produced is available online at pop.evemodel.org or is contained in the supplementary material in the manuscript.

https://pop.evemodel.org

## Data and Code Availability

Interactive web viewer and downloads for popEVE scores available at pop.evemodel.org. Code will be available at github.com/debbiemarkslab/popEVE.

## Acknowledgements

We thank all members of the Marks Lab and Dias & Frazer Group for valuable discussions. We would also like to thank Jack Nicoludis and Invitae for their assistance with training EVE models at scale. R.O, A.K, C.S, M.F, J.F, and D.S.M. are supported by a Chan Zuckerberg Initiative Award (Neurodegeneration Challenge Network, CZI2018-191853). H.S., and D.S.M. are supported by an NIH Transformational Research Award (TR01 1R01CA260415). C.S. is supported by the National Science Foundation Graduate Research Fellowship under Grant No. M.D and J.F. are supported by the Spanish Ministry of Science and Innovation (PID2022-140793NA-I00).

## Author Contributions

R.O., J.F., M.D. and D.S.M. conceived the end-to-end approach. R.O., J.F., M.D. built the models. R.O. and C.A.S. compiled and annotated the clinical and genomics data for analysis. A.W.K. supported model training. H.D.S., T.A.H., C.A.S, and R.O. performed the structural and functional analysis. T.A.H. developed the interactive web application. R.O., D.F., M.D., J.F., and D.S.M. wrote the manuscript. D.S.M., J.F., and M.D. led and supervised the project.

