## Supplementary material for "Proteome-wide model for human disease genetics": Marks - Extended Methods.pdf

### Supplementary Notes and Methods for Whole proteome disease variant prediction with deep generative modeling of evolutionary scale and population scale genetic variation

February 22, 2025

### 1 Methods

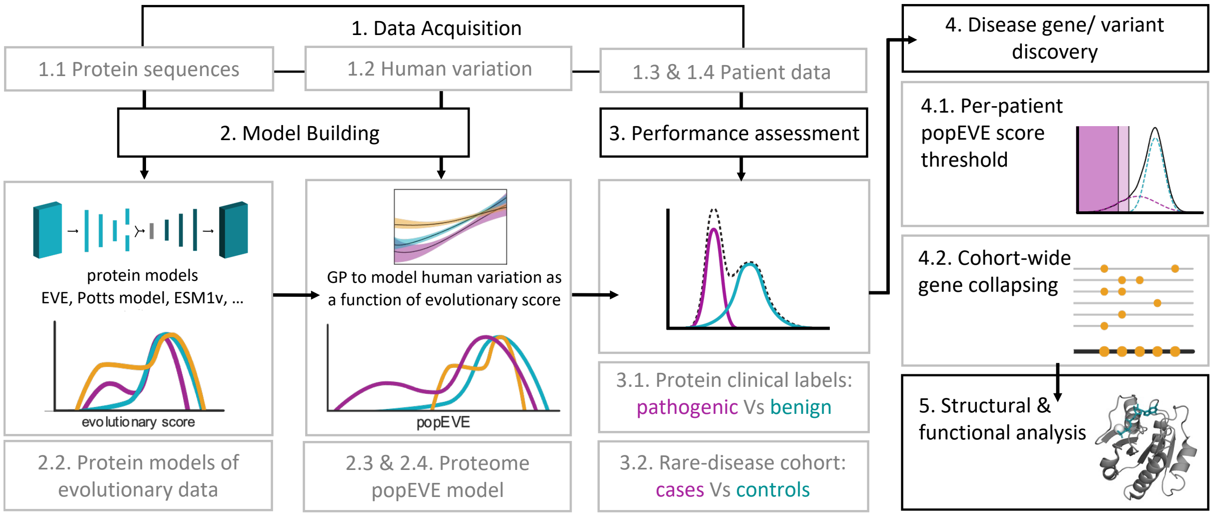

Structure of workflow and methods section

#### 1.1 Data acquisition

##### 1.1.1 Multiple sequence alignments

Described in section 1.2, some models require a preprocessing step, where training data is converted to multiple sequence alignments. Following a protocol similar to Hopf et al. [101] and Riesselman et al. [102], the EVCouplings pipeline [103], which builds on the profile HMM homology search tool Jackhmmer [104], was used to build multiple sequence alignments, where sequences were obtained from the UniRef100 database of non-redundant protein [105], downloaded in March 2022.

##### 1.1.2 Human variation data

Variants from UKBB 450k and from the final 500k release were annotated using VEP GRCh38 RefSeq and custom RefSeq annotation built from NCBI genebank files to maximize number of variants for pre-existing models. Variants were filtered for genotyping quality across all samples and annotations are filtered based off matching between RefSeq reference sequence and transcript sequences. When analyzing variants seen in the UKBB outside of training, we removed genes in which less than 95% of UK Biobank participants had at least 10x coverage [106].

##### 1.1.3 Developmental Disorder Cohorts

**Severe Developmental Disorder Metacohort** De novo mutations from a metacohort composed of subjects from the Deciphering Developmental Disorders study, GeneDx and Radboud

Medical center were aquired from Kaplanis et al. [107]. Quality filtering was performed by the respective centers as described in the supplement of Ref. [107]. The variants were re-annotated with VEP using GRCh37 RefSeq and custom mapping based on NCBI RefSeq assembly mapping files.

**Autism Spectrum and Unaffected Siblings Metacohort** De novo mutations from SFARI's SPARK and SCC cohorts and (the other two cohorts) were aquired from [108]. The variants were re-annotated with VEP using GRCh37 RefSeq and custom mapping based on NCBI RefSeq assembly mapping files. Sibling pairs with shared de novo variants were discarded due to the shear improbability of sharing DNMs between siblings, assumed to be some error either due to sequencing or the DNM calling pipeline.

**Deciphering Developmental Disorders Cohort** Variants from whole exome sequencing for the Deciphering Developmental Disorders cohort, a subset of the SDD metacohort, were re-annotated with VEP using GRCh37 RefSeq and custom mapping files. Variants were then filtered by quality based on the filters used in [107] to maximize the number of de novo variants seen (RD $> 7$, QD $\geq 3$, FS $\leq 60$, SOR $\leq 5$, $MQ \geq 50$, MQRankSum $> -2$, ReadPosRankSum $> -8$).

##### 1.1.4 ClinVar Benign and Pathogenic Variants

We made use of three sets of clinically labelled variants from the ClinVar public archive [109] for assessing the predictive performance of various models (described in Sec. 1.3.1). One set was downloaded in June 2023 and was processed with the same procedure used in Ref. [110]. Briefly, only missense variants with at least a one star rating were considered and class labels were remapped to a binary system where "benign" and "likely benign" were both relabeled as "Benign" and similarly "likely pathogenic" and "pathogenic" were relabelled as "Pathogenic". Two more sets of labels from 2019 and 2020, which were curated in Ref. [111] were also used for comparing the performance of supervised models. Results of this analysis can be found in Supplementary Figure 2.

##### 1.1.5 Deep mutational scans from ProteinGym

For assessing the predictive performance of a models based on their correlation with high-throughput functional assays, otherwise known as deep mutational scans, or multiplexed assays of variant effects, we consider the Human subset of Protein Gym [112] which are thought to be clinically relevant. Due to the method by which the reference sequences are chosen (Sec. 1.1.2), we do not have have sequence matches for all available assays. Thus the resulting test set consists of 23 assays across 18 proteins, so a modest expansion of the set considered in Ref. [110]. Results of this analysis can be found in Supplementary Figure 3.

#### 1.2 Model Building

##### 1.2.1 Overview of modelling strategy

From a methodological standpoint, our central goal is to rank the severity of the effect of genetic variants seen across an individual's proteome. To this end, we developed a probabilistic model which utilises protein sequence data from across diverse organisms (UniRef100) and within the human population (The UK Biobank). These two types of sequence data have complementary characteristics. As discussed in Ref. [110], sequence data from diverse organisms may be viewed as the result of millions of years of evolutionary experiments and looking for patterns of conservation in these sequences has long been appreciated as a powerful means of relating sequence to protein structure and function, enabling the relative effects of individual amino acid substitutions to be resolved. Sequence data from the human population on the other hand, and in particular, whole exome sequencing, provides a human specific means of measuring the degree of constraint acting on one coding region vs another. The intention in using both types of data, therefore, is to take advantage of the contrasting properties of these two data sets to achieve a model which can not only predict the effect of genetic variation across the proteome, but also do so with missense level resolution.

In the following sections, we first introduce the models used for identifying patterns of conservation across diverse organisms. We refer to these models as "evo" models. These models provide a "fitness" score for a given sequence of interest by obtaining an estimate for the log-odds

$$\sigma = \log\left(\frac{p(\boldsymbol{x})}{p(\boldsymbol{x}^{\text{ref}})}\right) , \tag{1}$$

where $\boldsymbol{x}$ represents the sequence of interest and $\boldsymbol{x}^{\text{ref}}$ is the reference sequence. In words, a sequence is considered "unfit" if it is an outlier compared to a reference sequence, with respect to the learned sequence distribution. We then introduce popEVE, a new model which takes as input these fitness scores and predicts the presence or absence of a variant in the UK Biobank, conditioned on the scores from these underlying models. This model naturally provides a new fitness score, which may be viewed as a rescaled/calibrated ensembling of the scores from the underlying models, such that the effect of variants in distinct proteins can be compared. We then describe our assessment of the performance of the model. We conclude that the model presents a significant improvement over previous models at ranking variants across genes and also provides a new state-of-the-art at ranking variants within the same gene, albeit a more modest step forwards.

##### 1.2.2 Modelling individual proteins using evolutionary data

It has recently emerged that unsupervised models which fit the distribution of sequences across diverse organisms can distinguish benign from pathogenic variants in genes already associated with disease, performing on par with high-throughput functional assays [110, 113]. This class of models have enjoyed rapid development, in part due to their diverse applications, spanning protein design, to pathogen forecasting [114–116]. We make use of two sub-classes of models which can be categorized as 1. alignment-based models; which use as training data the multiple sequence alignment obtained for a given gene of interest (Sec. 1.1.1), and 2. alignment-free models which are inspired by large language models and are trained on entire protein databases such as UniRef90. Here we summarise each model.

**The Bayesian Variational Autoencoder (EVE)** Variational autoencoders (VAEs) [117, 118] are a class of latent variable models which have been shown to be effective at capturing high-dimensional distributions in computer vision [119,120], natural language processing [121] and more. The assumption underlying a VAE is that the observed high-dimensional distribution is generated by a much smaller number of hidden (a.k.a latent) variables $z_i$. The generative story is thus

$$\boldsymbol{z} \sim \mathcal{N}(0, \boldsymbol{I}_D)$$
$$p(x_i^\alpha|\boldsymbol{z},\boldsymbol{\theta}) = \mathrm{softmax}((f^{\boldsymbol{\theta}}(\boldsymbol{z}))_i^\alpha)\,, \tag{2}$$

where $x_i^\alpha$ is an indicator function for the presence of amino acid $\alpha$ (superscript Greek indices) at position $i$ and the "decoder" $f^{\boldsymbol{\theta}}(\boldsymbol{z})$ is modelled with a fully-connected neural network, with spherical Gaussian prior for the parameters $\boldsymbol{\theta}$. In words, the VAE models the conditional probability of seeing the amino acid $\alpha$ at position $i$, given the latent variables $\boldsymbol{z}$. Parameter inference is achieved via the use of amortized inference, where we model the distribution $q(\boldsymbol{z}|x_{ij},\boldsymbol{\theta})$ with another fully-connect neural network, often referred to as the encoder. In Ref. [110] we found a symmetric relationship between between encoder and decoder to work well, with 3 layers, consisting of 2,000 - 1,000 - 300 and 300 - 1,000 - 2,000 nodes respectively.

To score a sequence, we use the evidence lower bound (ELBO) which is a lower bound on the log-marginal likelihood $p(\boldsymbol{x})$

$$\mathrm{ELBO}(\mathbf{x}) = N_{\text{eff}}\,.\,\mathbb{E}_{p(\boldsymbol{x})}\left[\mathbb{E}_{q(\theta_{\mathbf{p}}),q(\mathbf{z}|\mathbf{x})}\left(\log p(\boldsymbol{x}|\mathbf{z},\theta_{\mathbf{p}})\right) - D_{KL}(q(\mathbf{z}|\boldsymbol{x},\phi_{\mathbf{p}})||p(\mathbf{z}))\right] - D_{KL}(q(\theta_{\mathbf{p}})||p(\theta_{\mathbf{p}})) \tag{3}$$

where $N_{\text{eff}} = \sum_{n=1}^{N} w_{x_n}^{\mathrm{C}}$ and $w_{x_n}^{\mathrm{C}}$ is defined in Eq. (5). The fitness score is then simply

$$\sigma = \log\frac{p(\boldsymbol{x}|\theta_{\mathbf{p}})}{p(\boldsymbol{x}^{\text{ref}}|\theta_{\mathbf{p}})} \approx ELBO(\boldsymbol{x}) - ELBO(\boldsymbol{x}^{\text{ref}}) \tag{4}$$

**Sequence reweighting** All models used in this work make the false assumption that the training data is independently and identically distributed (iid). Ths iid assumption does not hold, due to phylogeny and also biases in which organisms have been sequenced. The fact that the VAE is trained on aligned data presents an opportunity to correct for these two biases with sequence re-weighting. Following the approach described in Ekeberg et al. [122], we re-weight each protein sequence $\boldsymbol{x}_i$ from a given MSA according to the reciprocal of the number of sequences in the corresponding MSA within a given Hamming distance cutoff $T$.

$$w_{\boldsymbol{x}_n}^{\mathrm{C}} = \left(\sum_{\substack{m=1\\m\neq n}}^{N} \mathbb{1}[\mathrm{Dist}(\boldsymbol{x}_n,\boldsymbol{x}_m) < T]\right)^{-1} \tag{5}$$

where $N$ is the number of sequences in the MSA, and bold, lowercase $\boldsymbol{x}$ represents a protein sequence, indexed by subscript Latin indices. As in Hopf et al. [101], we set $T = 0.2$ for all human proteins.

**Masked Language Model (ESM1v)** The transformer architecture has enabled the training of single, alignment free, models of essentially all known proteins. In this work we make use of ESM-1v [123,124], which is trained on UniRef90. It achieves a comparable performance to EVE on a number of tasks (see 1.3) and has complementary properties. By combining the two we expect to obtain a stronger model than would be achieved by building on EVE or ESM-1v alone.

ESM-1v [123,124] is a high-capacity 650M parameter language model which employs a form of self-supervision known as masking. During training, each sequence has a randomly sampled fraction of its amino acids replaced with a "mask" token and the network is then trained to predict the amino acids which have been masked. For each masked amino acid, the negative log likelihood of the missing amino acid, conditioned on the sequence context, is independently minimized.

$$\mathcal{L} = \mathbb{E}_{x\sim X}\mathbb{E}_M \sum_{i\in M} -\log p(x_i|x_{/M})\,. \tag{6}$$

Hence, in order for the model to successfully perform this task, the dependencies between the masked amino acid and the unmasked sequence context must be learned. k

###### 1.2.3 Estimating variant level constraint in humans using Gaussian Processes

While the models described in the previous section perform well at ranking variants within a given gene of interest, they do not perform well at ranking variants across genes (Figure 1, Supplementary Table 1). This is not surprising, since these models were not developed for that task and in the case of the alignment-based models, the models are trained completely independently for each coding region of interest. Indeed, to our knowledge, while proteome-wide predictions have been provided for numerous models (e.g. those in Fig 2e of main text), no model to date has been developed to explicitly address the problem of ranking variants across genes. As such, here we introduce what might be regarded as the first proteome model, which we call popEVE.

Similar to above, we define the evo score from one of the evo models, which we index $A$, with $A \in \{1,2\}$, as the log-odds between the sequence of interest $\boldsymbol{x}$ and some reference sequence $\boldsymbol{x}_{\text{ref}}$

$$\sigma^A = \log\left(\frac{p^A(\boldsymbol{x})}{p^A(\boldsymbol{x}^{\text{ref}})}\right)\,. \tag{7}$$

In what follows, sequences which differ from the reference sequence by a single amino acid substitution play a special role, so it is convenient to define $(\sigma_i^\alpha)_n^A$ as the score from model $A$ for a protein sequence, which differs from the reference $\boldsymbol{x}_n^{\text{ref}}$ sequence for protein $n$ solely by having amino acid $\alpha$ at position $i$.

We expect the probability of observing a sequence in the population to depend, in a fairly simple manner, on the score from the underlying evo models. We adopt a simple Bayesian, non-linear and non-parametric approach to modelling this relation; with the use of a Bernoulli likelihood, and latent Gaussian process. Specifically, we model the presence or absence of the variant in the UK Biobank as

$$p_n^A(y_i^\alpha|\sigma_{in}^{\alpha A}) = \text{Ber}\left(y_{in}^\alpha|\varphi(f_n^A(\sigma_{in}^{\alpha A}))\right) \tag{8}$$

where $y_{ijk} \in \{1, 0\}$ indicates the presence or absence of variant in the UK Biobank, the link function $\varphi(\cdot)$ is the inverse logit function (also referred to as the logistic function) $\varphi(z) = \exp(z)(1 + \exp(z))^{-1}$, and the function $f_n^A(\sigma)$ is drawn from a Gaussian process prior

$$f(\sigma) \sim GP(m(\sigma), \mathcal{K}(\sigma, \sigma')), \tag{9}$$

with zero mean function $m(\sigma) = 0$ and radial basis function kernel

$$\mathcal{K}(\sigma, \sigma') = \exp\left(-\gamma(\sigma - \sigma')^2\right). \tag{10}$$

The inferred function $f_n^A(\sigma)$ can be thought of as a new fitness score. The intuition is that by modelling the amount of variation seen per protein in the UK Biobank, $f_n^A(\sigma)$ essentially rescales the the evo score $\sigma_n^A$ to account for the degree of constraint acting on a per variant basis in the population, and how that constraint depends on $\sigma_n^A$, thus resulting in a score which can rank the pathogenicity of variants across different coding regions.

**Efficient function inference by restoring conjugacy with Pólya-Gamma data augmentation**
For each protein of interest, indexed $n$, and each underlying evo model, indexed $A$, we seek to infer the functions $f_n^A$. To do so, we consider the scores of all possible single amino acid substitutions in that protein and their corresponding labels $y_{in}^{\alpha}$, indicating if that variant has been observed, or not, in the UK Biobank. Dropping the indices $n$ and $A$ for compactness, we denote the training data as the set of scores $\boldsymbol{\sigma} = [\sigma_1^1, \ldots, \sigma_L^{19}] \in \mathbb{R}^N$ and $\boldsymbol{y} = [y_1, \ldots, y_N] \in \{0, 1\}^N$, where $L$ is the number of amino acids in the protein and $N = 19L$, being the total number of possible single amino acid substitutions. Let $\boldsymbol{f} = [f_1, \ldots, f_N]$, be the function values corresponding to the input $\boldsymbol{\sigma}$, then Eq. (8), together with the Gaussian process prior for $f$ implies

$$p(\boldsymbol{f}|\boldsymbol{y}, \boldsymbol{\sigma}) \propto p(\boldsymbol{y}|\boldsymbol{f})p(\boldsymbol{f}|\boldsymbol{\sigma})\,, \tag{11}$$

where $p(\boldsymbol{f}|\boldsymbol{\sigma}) = \mathcal{N}(\boldsymbol{f}|0, K_{NN})$, with $K_{NN}$ denoting the kernel matrix evaluated at the training points. Thus, in contrast to models with a Gaussian process prior and Gaussian likelihood, inference is analytically intractable due to the Gaussian prior not being conjugate to the Bernoulli likelihood.

One appealing approach to overcoming this is to introduce additional latent variables that restore conjugacy. Following Ref. [125], we introduce the auxiliary variables $\boldsymbol{\omega}$ and define the augmented likelihood to factorise as

$$p(\boldsymbol{y}, \boldsymbol{\omega}) = p(\boldsymbol{y}|\boldsymbol{f}, \boldsymbol{\omega})p(\boldsymbol{\omega}) \tag{12}$$

The goal then is to find a prior $p(\omega)$ to satisfy two properties: 1. That when marginalising out $\omega$ the original model is recovered. 2. The Gaussian prior $p(\boldsymbol{f})$ is conjugate to the likelihood $p(\boldsymbol{y}|\boldsymbol{f}, \boldsymbol{\omega})$. These conditions are satisfied by the Pólya-gamma distribution, which may be thought of as an infinite convolution of Gamma distributions. *i.e.* $\omega \sim \mathrm{PG}(\mathrm{b}, \mathrm{c})$, where

$$\omega = \frac{1}{2\pi^2} \sum_{m=1}^{\infty} \frac{g_m}{(m - \frac{1}{2})^2 + (\frac{c}{2\pi})^2}\,, \tag{13}$$

and $g_m \sim \Gamma(b, 1)$. Alternatively, we can define the Pólya-gamma distribution in terms of its moment generating function (These two definitions are related by the Laplace transform):

$$\mathbb{E}_{\mathrm{PG}}[\exp(-\omega t)] = \frac{1}{\cosh^b\left(\sqrt{\frac{t}{2}}\right)}. \tag{14}$$

This second definition is useful, since it suggests that the logistic link function may be expressed in terms of Pólya-gamma variables

$$\begin{aligned}\varphi(z_i) &= \frac{\exp(z_i)}{(1+\exp(z_i))} \\ &= \frac{\exp(\frac{z_i}{2})}{2\cosh(\frac{z_i}{2})} \\ &= \frac{1}{2}\int \exp\left(\frac{z_i}{2} - \frac{z_i^2}{2}\omega_i\right) p(\omega_i)\mathrm{d}\omega_\mathrm{i}\,.\end{aligned} \tag{15}$$

Hence, substituting $z_i = y_i f(\sigma_i)$, we obtain

$$p(\boldsymbol{y}|\boldsymbol{\omega}, \boldsymbol{f}) \propto \exp\left(\frac{1}{2}\boldsymbol{y}^\top \boldsymbol{f} - \frac{1}{2}\boldsymbol{f}^\top \Omega \boldsymbol{f}\right), \tag{16}$$

where $\Omega = \mathrm{diag}(\boldsymbol{\omega})$ is the diagonal matrix of the Pólya-gamma variables. This augmented likelihood is conjugate to $p(\boldsymbol{f})$ as required.

Developed in [126], once conditional conjugacy is restored, it is possible to derive closed-form updates for variational inference with natural gradients and a learning rate close to one, enabling highly efficient inference of $\boldsymbol{f}$.

**Making the models scale with inducing points** Inference in GPs with Gaussian a likelihood, while exact, take $\mathcal{O}(N^3)$ time and hence additional methods are required in order to perform inference when the training data is large. One such method is to learn a "summary" of the data with $M \ll N$ pseudo inputs, otherwise known as inducing points [127] and hence reduce the complexity to $\mathcal{O}(M^3)$. Following [126], we introduce $M$ additional variable $\boldsymbol{u} = [u_1, \ldots, u_M]$, where the function values of the GP $\boldsymbol{f}$ are related to $\boldsymbol{u}$ by

$$\begin{aligned}p(\boldsymbol{f}|\boldsymbol{u}) &= \mathcal{N}(\boldsymbol{f}|K_{NM}K_{MM}^{-1}\boldsymbol{u}, \tilde{K}) \\ p(\boldsymbol{u}) &= \mathcal{N}(\boldsymbol{u}|0, K_{MM}),\end{aligned} \tag{17}$$

where $k_{MM}$ is the kernel matrix resulting from evaluating the kernel at the $M$ inducing points, $K_{NM}$ is the kernel between the training points and the inducing points and $\tilde{K} = K_{NN} - K_{NM}K_{MM}^{-1}K_{MN}$.

Hence, the complete joint distribution of our model is given by

$$p(\boldsymbol{y}, \boldsymbol{\omega}, \boldsymbol{f}, \boldsymbol{u}) = p(\boldsymbol{y}|\boldsymbol{\omega}, \boldsymbol{f})p(\boldsymbol{\omega})p(\boldsymbol{f}|\boldsymbol{u})p(\boldsymbol{u})\,. \tag{18}$$

**Implementation** This model is implemented in GPyTorch [128] and will be made publicly available via a dedicated GitHub repository soon.

##### 1.2.4 Ensembles of models with only partially intersecting domains

Ensembles of models can often achieve a performance similar to, and sometimes even stronger than, the strongest constituent model [129]. Our setup provides a novel opportunity to build a highly performant ensemble model by incorporating the scores from multiple evo models. By training a separate Gaussian process model for each evo model, we naturally create directly comparable scores between models, thereby enabling the typical, but potentially problematic, standardization step to be bypassed entirely. We also gain an additional measure of uncertainty since we can assess the degree of concordance between the models. We define the popEVE score $\bar{\sigma}$ to simply be the mean of the means of the posteriors of each GP, for each evo model who's domain contains the variant of interest.

$$\bar{\sigma}_{ij}^{\alpha} = \frac{1}{Q_{ij}^{A}} \sum_{A=1}^{Q_{ij}^{\alpha}} \mathbb{E}\left[f_j^A(\sigma_i^{\alpha})\right] , \tag{19}$$

where $Q_{ij}^{A}$ is the number of evo models capable of making a prediction for the amino acid substitution $\alpha$ at position $i$ in protein $j$.

An alternative approach would have been to ensemble the scores from the evo models directly, and then only train a single GP per protein, rather than one per evo model, per protein. Choosing the latter brings a number of advantages. First, the scores from the underlying evo models do not have the same distribution, meaning that some form of standardization of the scores would be required if they were to be ensembled directly. In contrast, the function scores $f_j^A$ are directly comparable, thereby providing a principled basis for ensembling (and uncertainty estimation. This point is particularly important for protein modelling, since the domain of each evo model can be different, hence the number of models contributing to scoring a given amino acid substitution can vary. This incomplete overlap of coverage between the models leads to discontinuities in the ensembled score, potentially damaging the performance of the model. This problem is to some degree alleviated with the use of the GP scores for ensembling. Second, by training independent GPs for each evo model, it facilitates future model updates, where additional evo models may be included in a way that is flexible with regard to reweighting the relative importance of each model (*i.e.* using a weighted sum in Eq. (1.2.4)), which will become increasingly necessary as more models are included, where some are more similar than others.

#### 1.3 Performance Assessments

We constructed a number of tests in order to assess key properties of the model, and compare its performance to preexisting models. Broadly, we wanted to assess the model's ability to perform two tasks; The ability to rank the pathogencity of variants within the same gene and across the proteome. The former is especially important for burden testing analysis (Sec. 1.4.2) and analysis at the level of individual proteins more generally, while the latter is critical for identifying candidate disease causing variants in whole exome data and the discovery of disease causing variants in genes where no disease association is currently known. Given that the popEVE score for each constituent model is simply a one-dimensional, often monotonic, function of the underlying score $f_n^A(\sigma)$, we expect the performance, on a per protein basis, to only be a modest improvement over the underlying evo models. This is because the ranking of variants will be approximately preserved, so the only potential benefit of the new score comes from the ensembling aspect of the model. In contrast, when comparing variants across proteins, popEVE should provide a significant improvement as compared to the underlying models.

##### 1.3.1 Comparing model performance within proteins

To assess model performance at ranking the pathogenicity of variants within the same gene, similar to Ref. [110] we consider two tests – correlation of model predictions with deep mutational scans (multiplexed assays of variant effects) (Sec. 1.1.5) and ability to predict benign and pathogenic labels in ClinVar (Sec. 1.1.1).

To assess concordance with deep mutational scans, we compute the Spearman's correlation between the model score and the experimental readout (*e.g.* expression). Supplementary Figure 3 shows a comparison of performance of popEVE and its constituent evo models (described in Sec. 1.2.2) (Top) and other state-of-the-art models designed to predict pathogenic variants (Bottom), which were downloaded from dbNSFP version 4.7 [130] and are the same set of models that were analysed in Ref. [111]. On average, popEVE outperforms all models as well as having more consistent performance; typically ranking top, or near the top, for each protein, as expected given that it is an ensemble model (described in Sec 1.2.4).

For comparison with clinical labels, we made use of three curated sets of ClinVar labels (Sec. 1.1.4). For assessing the performance of popEVE and its constituents, none of which make use of clinical labels during training, we used the 2023 set of clinical labels following the same procedure as in Ref. [110]. This set enables performance to be assessed on a per-protein basis across 860 proteins. For comparison with other state-of-the-art models, designed to predict pathogenic variants, most of which use supervised learning on clinical labels, we use the recently released ClinGen curated 2019 and 2020 sets, where labels used in training by these models have been removed [111]. Requiring a gene to have at least 5 Benign and 5 Pathogenic labels to be included in the benchmark, with these sets we were able to assess the performance across 50 and 31 proteins and in 2019 and 2020 data sets respectively. Comparing popEVE to its constituent models (Supplementary Table 1), we see that on average, popEVE outperforms both EVE and ESM-1v. We also find that on average popEVE matches (2019 set, Supplementary Figure 2a, Supplementary Table 1), or outperforms (2020 set, Supplementary Figure 2b, Supplementary Table 1) all current state-of-the-art models designed to predict pathogenic variants.

##### 1.3.2 Comparing model performance across proteins

To assess the performance of the model at ranking the pathogenicity of variants across proteins we constructed a test set based on the de novo variants found in patients with severe developmental disorders (Sec 1.1.3) and the unaffected siblings of patients with autism spectrum disorder 1.1.3. The basic objective was to test if the model could distinguish patients harbouring a de novo variant likely causal for a severe developmental disorder from patients whose de novo variants are not expected to play a role in disease. To this end, we built approximately balanced "case" and "control" sets. To obtain the case set we only considered individuals who had at least one de novo variant in a gene known to be involved in a developmental disorder, according to DDG2P [131], downloaded January 2023. This gives us a "diagnosed" subset of cases, in line with Kaplanis et al [107]. However, not all of these individuals are expected to have a pathogenic de novo missense variant. As such, we adjust our recall to an expected maximum recall - given the background mutation rate, we see an excess of 2982 de novo misense variants in the 5625 cases in this subset of the cohort so we expect 2982/5625=53% of these individuals to have a pathogenic de novo missense variant. We rescaled the recall to reflect this. The resulting test set is provided in Supplementary Table 3. Shown in Figure 2f of the main text and Supplementary Figure 9a, we find popEVE to outperform all current state-of-the-art models designed to predict pathogenic variants, as well as its constituent evo models (Supplementary Table 1).

We then constructed a test to assess ability to recover severe developmental disorder cases either from de novo mutations alone or with the entirety of their whole exome sequencing without overpredicting pathogenicity in the general population. For each model, we segmented the scores into score thresholds and for each threshold we calculated the percent of individuals in the UK biobank with a variant as or more pathogenic than that value. Then we calculated the percent of individuals in the SDD meta-cohort or DDD sub-cohort with a variant as or more pathogenic (Figure 2d & 3e, Supplemental Figure 9 d & f). These plots can be thought of as a direct comparison between two cumulative distributions for developmental disorder cases and UK biobank healthy controls. For the the SDD meta-cohort, we selected those with a de novo missense variant in a gene previously discovered in the same cohort by DeNovoWEST [107], making this a test of high-confidence "diagnosed" individuals.

Next, we tested how well models distinguished de novo mutations from inherited mutations in the DDD sub-cohort. For each score threshold, we took the cases with a de novo mutation as or more pathogenic and computed the percent of these cases that had their de novo mutation ranked more pathogenic than their inherited variants (Figure 2b & Supplementary Figure 9f). For example, at a score of -5.056 (our severe threshold), 513 DDD individuals have a de novo missense variant that or more pathogenic, and, in 98% of these cases, their de novo variant is ranked their most pathogenic variant. The rate of decay shows each model's ability to rank likely causal variants as the most pathogenic based on each score threshold.

#### 1.4 Analysis of patient data

We explored two approaches to analysing de novo mutations from a metacohort composed of subjects from the Deciphering Developmental Disorders study, GeneDx and Radboud Medical center in the hope that popEVE may provide novel evidence for the genetic diagnosis of currently unsolved cases. One approach was burden testing, which has the important advantage of gaining statistical power thanks to the design of the cohort. The second approach was analyse patient data on a case by case basis. The advantage of this second approach being that we may be able to find evidence for the genetic diagnosis of diseases which are so rare that building a sufficiently large cohort to discover via burden testing is not possible. The second approach is also of course more widely applicable albeit less robust.

##### 1.4.1 Direct case-variant association

Based on the set of de novo variants from cases and controls, described in Sec. 1.3.2 we constructed a Bayesian Gaussian Mixture Model to determine a score cutoff as

$$\begin{aligned}
&\boldsymbol{\mu}_1 \sim \mathcal{N}(\boldsymbol{\mu}_0, \boldsymbol{\Sigma}_0) \\
&\boldsymbol{\mu}_2 \sim \mathcal{N}(\boldsymbol{\mu}_0, \boldsymbol{\Sigma}_0) \\
&\lambda_1 \sim \text{Lognormal}(\mu_\lambda, \sigma_\lambda) \\
&\lambda_2 \sim \text{Lognormal}(\mu_\lambda, \sigma_\lambda) \\
&\boldsymbol{\pi} \sim \text{Dirichlet}(\boldsymbol{\alpha}) \\
\text{For } i = 1, \ldots, N: \\
&a_i \sim \text{Categorical}(\boldsymbol{\pi}) \\
&\mathbf{x}_i \sim \mathcal{N}(\boldsymbol{\mu}_{a_i}, \lambda_{a_i})
\end{aligned} \tag{20}$$

where $\boldsymbol{\mu}_0 = -3.6$ and $\boldsymbol{\Sigma}_0 = 0.7$. We then identified an uncertainty cutoff corresponding to a greater than 99.99% likelihood of being in the lower fitness distribution, $\bar{\sigma} \leq -5.056$. We found that constructing the model with solely the de novo variants from the cases resulted in a similar threshold.

Based on the threshold $\bar{\sigma} \leq -5.056$, we then searched the full developmental disorder cohort (Sec. 1.1.3) for individuals with at least one de novo variant below this score, who also do not have any loss of function variants. For those individuals, we consider these variants to be strong candidates for being the causal variant. In addition to the high accuracy of the Gaussian mixture model, further support for variants below this threshold is provided by noting the high enrichment of such variants in the metacohort compared to the naive expectation (Figure 2d of main text). A full list of the variants found in the metacohort together with their associated popEVE scores may be found in Supplementary Table 3.

##### 1.4.2 Gene-collapsing model

For comparison with previous methods, we built gene-collapsing models to identify gene-disorder associations in the SDD cohort in the style of DeNovoWEST [132]. The probability of seeing an overall score worse for a particular gene than the observed score $x_{obs}$ is assessed by testing from 0 to N until the likelihood of the number of mutations, modeled using a Poisson distribution and mutation rates (described below) goes to zero. De novo mutation rates of bases in their genomic context, from Samocha et al 2016, are used to both determine the number of expected de novo mutations in a particular gene of a given cohort size and to select mutations when performing simulations. For each gene, we performed 10,000 simulations of gene mutations expected to be seen in a single generation of a cohort this size. In addition to testing the overall enrichment and likelihood of the observed score $x_{obs}$ given the distribution of scores in a single gene, we added a component that assesses the likelihood of seeing the observed score across the entire proteome.

$$p(\text{gene}) = \sum_{n=0}^{N} P(n = n_{gene}|\lambda_{gene}) P(x_{gene} \leq x_{obs}|n) P(x_{proteome} \leq x_{obs}|n) \tag{21}$$

We selected our significance cutoff by dividing 0.05 by the total number of tests (or total number of genes/proteins we have modeled) - p<0.05/18,395.

#### 1.5 Structural and Functional Analysis of deleterious mutations

##### 1.5.1 Functional Similarity of Novel and Known DD Genes

We compared the functional properties of our novel genes in comparison to known developmental disorder genes [131] across features known to differentiate DD-genes from genes not associated with developmental disorders similar to Kaplanais et al 2020 [107]. We calculated the enrichment of these variables in either our novel genes or known DD-genes as compared to non-DD genes. Results of this analysis are in Supplementary Figure 10, with full gene annotations for all features found in Supplementary Table 6 and 7.

- · Interactions: We selected protein-protein interactions (BioGRID [133]) enriched in known DD-genes compared to non-DD genes. To ensure these interactions are generalizable, we selected those that are significantly enriched in known DD-genes (p<0.05 after Benjamini-Hochberg correction) and present in at least 10% of known DD-genes (n>223).
- · Median Fetal RPKM: Expression in fetal brain, median RPKM across samples, from Allen Brain Atlas [134].
- · Relevant GO terms MF (Molecular Function) and BP (Biological Processes): We selected gene ontology (GO) terms enriched in known DD-genes compared to non-DD genes using DAVID [135]. To ensure these terms are generalizable, we selected those that are significantly enriched in known DD-genes (p<0.05 after Benjamini-Hochberg correction) and present in at least 10% of known DD-genes (n>223).
- · Haploinsufficient: a binary variable of genes with evidence of haploinsufficiency (dosage pathogenicity level 3) from the ClinGen Dosage Sensitivity Map [136].
- · Human Essential: a binary variable of whether a gene was deemed essential in human cell lines based on CRISPR screens [137].
- · ACMG Genes: a binary variable of whether a gene is a clinically actionable gene according to the American College of Medical Genetics and Genomics v2.0 [138].
- · Somatic Drivers: a binary variable of whether the gene is known to be a somatic driver gene [139].
- · Mouse Essential: a binary variable of whether a gene is essential in mice, i.e. homozygous knockouts of that gene resulted in lethality [140] [141].
- · LoF Tolerant: a binary variable of whether a gene is tolerant of homozygous LoF mutations in humans [142].

##### 1.5.2 Functional Network

We created a functional gene network with 130 de novo novel genes from popEVE (with a 99.99 threshold) and significant genes from DeNovoWEST [132] using STRING [143] as shown in Supplementary Figure 11 and Supplementary Table 8. We show edges from medium-confidence (0.4) experiments. Genes were denoted as previously discovered if they were already observed in DDG2P [131] or DeNovoWEST.

Additionally, we used medium-confidence (0.4) experiments annotations to calculate the average node degree of DDG2P genes and DeNovoWEST genes with and without our significant popEVE genes included, both from de novo analysis and no trios analysis with results in Supplementary Figure 12. In order to see the relative difference that the significant genes make vs a random set of genes, we performed t-tests 100,000 times with random samples of genes from the whole human genome (with the known and popEVE genes excluded).

##### 1.5.3 Manual Structure Analysis

From the 131 novel popEVE mutations, we individually investigated structures for the top 20 most predicted deleterious. We only analyzed cryo-electron microscopy or crystallographic structures where our mutation is included in the resolved protein structure. To enhance our analysis, we prioritized structures that exhibited interactions with other proteins and/or ligands. This allowed us to capture and understand the potential consequences of these interactions. The structural figures were generated using PyMol [144]. The mutated residues were depicted as sticks, with their corresponding elemental colors assigned for clarity. All distances listed were calculated with the distance function in PyMol.

##### 1.5.4 Comparative Analysis of Protein-Ligand Interactions Using a Null Model

Taking de novo variants from the SDD metacohort and inherited variants from the DDD subset, we conducted a comparative interaction analysis for these sites compared to a null model. The null model calculated the distance between every position on the variant-containing chain and the closest ligand to the mutant of interest. Protein structures with resolved crystal structures were programmatically processed using the Evcouplings PDB reader, and distance calculations were determined using the Evcouplings compare package [145]. Z scores were calculated by subtracting the average distance between every position on the chain (excluding the variant position) from the distance between the variant position on the chain and the ligand. Variants were filtered before Z score calculation if the percentage difference between the reported protein chain length and the full protein length was less than 2 standard deviations from the mean. Full Results are in Supplementary Table 9.

##### 1.5.5 Functional interactions in 3D structures for high-scoring pathogenic variants

Available 3D structures for proteins with high-scoring pathogenic variants were retrieved and mapped to the target sequence by alignment-based search against sequences in the SIFTS database [146] using the EVcouplings package [145] with a single iteration of HMMER [147] at an inclusion threshold of 0.2 bits/residue. High-scoring pathogenic variants were considered to have positive evidence for functional interactions if there was at least one 3D contact between the variant position and another non-self PDB entity within a minimum atom distance of 8Å in at least of the identified structures, excluding waters and the most frequent chemical additives for structure determination (entity information obtained from the PDBe entry REST API [148]).

[109] Melissa J Landrum, Jennifer M Lee, Mark Benson, Garth R Brown, Chen Chao, Shanmuga Chitipiralla, Baoshan Gu, Jennifer Hart, Douglas Hoffman, Wonhee Jang, Karen Karapetyan, Kenneth Katz, Chunlei Liu, Zenith Maddipatla, Adriana Malheiro, Kurt McDaniel, Michael Ovetsky, George Riley, George Zhou, J Bradley Holmes, Brandi L Kattman,

and Donna R Maglott. ClinVar: improving access to variant interpretations and supporting evidence. *Nucleic Acids Research*, 46(D1):D1062–D1067, January 2018.

[110] Jonathan Frazer, Pascal Notin, Mafalda Dias, Aidan Gomez, Joseph K Min, Kelly Brock, Yarin Gal, and Debora S Marks. Disease variant prediction with deep generative models of evolutionary data. *Nature*, 599(7883):91–95, 2021.

[111] Vikas Pejaver, Alicia B Byrne, Bing-Jian Feng, Kymberleigh A Pagel, Sean D Mooney, Rachel Karchin, Anne O'Donnell-Luria, Steven M Harrison, Sean V Tavtigian, Marc S Greenblatt, et al. Calibration of computational tools for missense variant pathogenicity classification and clingen recommendations for pp3/bp4 criteria. *The American Journal of Human Genetics*, 109(12):2163–2177, 2022.

[112] Pascal Notin, Mafalda Dias, Jonathan Frazer, Javier Marchena Hurtado, Aidan N Gomez, Debora Marks, and Yarin Gal. Tranception: protein fitness prediction with autoregressive transformers and inference-time retrieval. In *International Conference on Machine Learning*, pages 16990–17017. PMLR, 2022.

[113] Benjamin J Livesey and Joseph A Marsh. Updated benchmarking of variant effect predictors using deep mutational scanning. *Molecular Systems Biology*, page e11474.

[114] Jung-Eun Shin, Adam J Riesselman, Aaron W Kollasch, Conor McMahon, Elana Simon, Chris Sander, Aashish Manglik, Andrew C Kruse, and Debora S Marks. Protein design and variant prediction using autoregressive generative models. *Nature communications*, 12(1):2403, 2021.

[115] Nicole N Thadani, Sarah Gurev, Pascal Notin, Noor Youssef, Nathan J Rollins, Daniel Ritter, Chris Sander, Yarin Gal, and Debora S Marks. Learning from prepandemic data to forecast viral escape. *Nature*, pages 1–8, 2023.

[116] Zachary Wu, Kadina E Johnston, Frances H Arnold, and Kevin K Yang. Protein sequence design with deep generative models. *Current opinion in chemical biology*, 65:18–27, 2021.

[117] Diederik P Kingma and Max Welling. Auto-encoding variational bayes, 2014.

[118] Danilo Jimenez Rezende, Shakir Mohamed, and Daan Wierstra. Stochastic backpropagation and approximate inference in deep generative models, 2014.

[119] Arash Vahdat and Jan Kautz. Nvae: A deep hierarchical variational autoencoder. *arXiv preprint arXiv:2007.03898*, 2020.

[120] Aditya Ramesh, Mikhail Pavlov, Gabriel Goh, Scott Gray, Chelsea Voss, Alec Radford, Mark Chen, and Ilya Sutskever. Zero-shot text-to-image generation. *arXiv preprint arXiv:2102.12092*, 2021.

[121] Samuel R. Bowman, Luke Vilnis, Oriol Vinyals, Andrew M. Dai, Rafal Jozefowicz, and Samy Bengio. Generating sentences from a continuous space, 2016.

[122] Magnus Ekeberg, Cecilia Lövkvist, Yueheng Lan, Martin Weigt, and Erik Aurell. Improved contact prediction in proteins: Using pseudolikelihoods to infer Potts models. *Physical Review E*, 87(1), January 2013.

[123] Joshua Meier, Roshan Rao, Robert Verkuil, Jason Liu, Tom Sercu, and Alex Rives. Language models enable zero-shot prediction of the effects of mutations on protein function. *Advances in Neural Information Processing Systems*, 34:29287–29303, 2021.

[124] Alexander Rives, Siddharth Goyal, Joshua Meier, Demi Guo, Myle Ott, C. Lawrence Zitnick, Jerry Ma, and Rob Fergus. Biological structure and function emerge from scaling unsupervised learning to 250 million protein sequences. preprint, Synthetic Biology, April 2019.

[125] Nicholas G Polson, James G Scott, and Jesse Windle. Bayesian inference for logistic models using pólya–gamma latent variables. *Journal of the American statistical Association*, 108(504):1339–1349, 2013.

[126] Florian Wenzel, Théo Galy-Fajou, Christan Donner, Marius Kloft, and Manfred Opper. Efficient gaussian process classification using pòlya-gamma data augmentation. In *Proceedings of the AAAI Conference on Artificial Intelligence*, volume 33, pages 5417–5424, 2019.

[127] Edward Snelson and Zoubin Ghahramani. Sparse gaussian processes using pseudo-inputs. *Advances in neural information processing systems*, 18, 2005.

[128] Jacob R Gardner, Geoff Pleiss, David Bindel, Kilian Q Weinberger, and Andrew Gordon Wilson. Gpytorch: Blackbox matrix-matrix gaussian process inference with gpu acceleration. In *Advances in Neural Information Processing Systems*, 2018.

[129] Zhi-Hua Zhou. *Ensemble methods: foundations and algorithms*. CRC press, 2012.

[135] Brad T Sherman, Ming Hao, Ju Qiu, Xiaoli Jiao, Michael W Baseler, H Clifford Lane, Tomozumi Imamichi, and Weizhong Chang. David: a web server for functional enrichment analysis and functional annotation of gene lists (2021 update). *Nucleic acids research*, 50(W1):W216–W221, 2022.

[136] Heidi L Rehm, Jonathan S Berg, Lisa D Brooks, Carlos D Bustamante, James P Evans, Melissa J Landrum, David H Ledbetter, Donna R Maglott, Christa Lese Martin, Robert L Nussbaum, et al. Clingen—the clinical genome resource. *New England Journal of Medicine*, 372(23):2235–2242, 2015.

[137] Traver Hart, Amy Hin Yan Tong, Katie Chan, Jolanda Van Leeuwen, Ashwin Seetharaman, Michael Aregger, Megha Chandrashekhar, Nicole Hustedt, Sahil Seth, Avery Noonan, et al. Evaluation and design of genome-wide crispr/spcas9 knockout screens. *G3: Genes, Genomes, Genetics*, 7(8):2719–2727, 2017.

[138] Sarah S Kalia, Kathy Adelman, Sherri J Bale, Wendy K Chung, Christine Eng, James P Evans, Gail E Herman, Sophia B Hufnagel, Teri E Klein, Bruce R Korf, et al. Recommendations for reporting of secondary findings in clinical exome and genome sequencing, 2016 update (acmg sf v2. 0): a policy statement of the american college of medical genetics and genomics. *Genetics in medicine*, 19(2):249–255, 2017.

[139] Iñigo Martincorena, Keiran M Raine, Moritz Gerstung, Kevin J Dawson, Kerstin Haase, Peter Van Loo, Helen Davies, Michael R Stratton, and Peter J Campbell. Universal patterns of selection in cancer and somatic tissues. *Cell*, 171(5):1029–1041, 2017.

[140] Judith A Blake, Carol J Bult, James A Kadin, Joel E Richardson, Janan T Eppig, and Mouse Genome Database Group. The mouse genome database (mgd): premier model organism resource for mammalian genomics and genetics. *Nucleic acids research*, 39(suppl_1):D842–D848, 2010.

[141] Benjamin Georgi, Benjamin F Voight, and Maja Bućan. From mouse to human: evolutionary genomics analysis of human orthologs of essential genes. *PLoS genetics*, 9(5):e1003484, 2013.

[142] Genome Aggregation Database Consortium, Konrad J. Karczewski, Laurent C. Francioli, Grace Tiao, Beryl B. Cummings, Jessica Alföldi, Qingbo Wang, Ryan L. Collins, Kristen M. Laricchia, Andrea Ganna, Daniel P. Birnbaum, Laura D. Gauthier, Harrison Brand, Matthew Solomonson, Nicholas A. Watts, Daniel Rhodes, Moriel Singer-Berk, Eleina M. England, Eleanor G. Seaby, Jack A. Kosmicki, Raymond K. Walters, Katherine Tashman, Yossi Farjoun, Eric Banks, Timothy Poterba, Arcturus Wang, Cotton Seed, Nicola Whiffin, Jessica X. Chong, Kaitlin E. Samocha, Emma Pierce-Hoffman, Zachary Zappala, Anne H. O'Donnell-Luria, Eric Vallabh Minikel, Ben Weisburd, Monkol Lek, James S. Ware, Christopher Vittal, Irina M. Armean, Louis Bergelson, Kristian Cibulskis,

Kristen M. Connolly, Miguel Covarrubias, Stacey Donnelly, Steven Ferriera, Stacey Gabriel, Jeff Gentry, Namrata Gupta, Thibault Jeandet, Diane Kaplan, Christopher Llanwarne, Ruchi Munshi, Sam Novod, Nikelle Petrillo, David Roazen, Valentin Ruano-Rubio, Andrea Saltzman, Molly Schleicher, Jose Soto, Kathleen Tibbetts, Charlotte Tolonen, Gordon Wade, Michael E. Talkowski, Benjamin M. Neale, Mark J. Daly, and Daniel G. MacArthur. The mutational constraint spectrum quantified from variation in 141,456 humans. *Nature*, 581(7809):434–443, May 2020.

[143] Damian Szklarczyk, Rebecca Kirsch, Mikaela Koutrouli, Katerina Nastou, Farrokh Mehryary, Radja Hachilif, Annika L Gable, Tao Fang, Nadezhda T Doncheva, Sampo Pyysalo, Peer Bork, Lars J Jensen, and Christian von Mering. The STRING database in 2023: protein–protein association networks and functional enrichment analyses for any sequenced genome of interest. *Nucleic Acids Research*, 51(D1):D638–D646, 11 2022.

[144] Schrödinger, LLC. The PyMOL molecular graphics system, version 1.8. November 2015.

[145] Thomas A Hopf, Anna G Green, Benjamin Schubert, Sophia Mersmann, Charlotta P I Schärfe, John B Ingraham, Agnes Toth-Petroczy, Kelly Brock, Adam J Riesselman, Perry Palmedo, Chan Kang, Robert Sheridan, Eli J Draizen, Christian Dallago, Chris Sander, and Debora S Marks. The EVcouplings python framework for coevolutionary sequence analysis. *Bioinformatics*, 35(9):1582–1584, May 2019.

[146] Sameer Velankar, José M Dana, Julius Jacobsen, Glen van Ginkel, Paul J Gane, Jie Luo, Thomas J Oldfield, Claire O'Donovan, Maria-Jesus Martin, and Gerard J Kleywegt. SIFTS: Structure integration with function, taxonomy and sequences resource. *Nucleic Acids Res.*, 41(Database issue):D483–9, January 2013.

[147] Sean R Eddy. Accelerated profile HMM searches. *PLoS Comput. Biol.*, 7(10):e1002195, October 2011.

[148] David R Armstrong, John M Berrisford, Matthew J Conroy, Aleksandras Gutmanas, Stephen Anyango, Preeti Choudhary, Alice R Clark, Jose M Dana, Mandar Deshpande, Roisin Dunlop, Paul Gane, Romana Gáborová, Deepti Gupta, Pauline Haslam, Jaroslav Koča, Lora Mak, Saqib Mir, Abhik Mukhopadhyay, Nurul Nadzirin, Sreenath Nair, Typhaine Paysan-Lafosse, Lukas Pravda, David Sehnal, Osman Salih, Oliver Smart, James Tolchard, Mihaly Varadi, Radka Svobodova-Vařeková, Hossam Zaki, Gerard J Kleywegt, and Sameer Velankar. PDBe: improved findability of macromolecular structure data in the PDB. *Nucleic Acids Res.*, 48(D1):D335–D343, January 2020.
