## Supplementary material for "Proteome-wide model for human disease genetics": Marks - Supplementary Figures.pdf

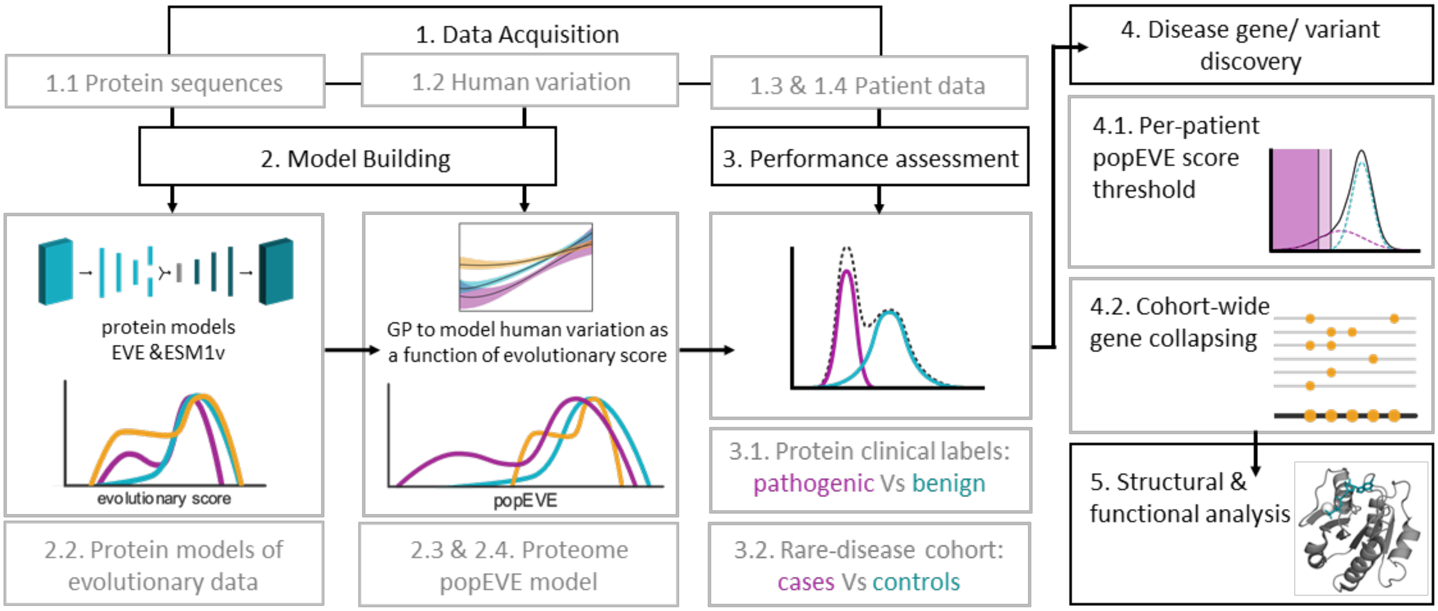


**Supplementary Figure 1. Structure of workflow and methods section.**
Our workflow for this project involved 1. data acquisition, 2. model building, 3. validation and performance assessment, 4. analysis of patient data towards discovery of disease variants and genes, and 5. structural and functional analysis of candidate disease variants. Model building has two main components: modeling of protein sequences across the entire tree of life from which we obtain a protein level fitness score and predicting the presence of a variant in the UKbiobank given its fitness score using Gaussian Processes, from which we obtain a score for the spectrum of pathogenicity across the human proteome, the popEVE score. We assess the performance of popEVE at predicting pathogenicity within proteins – by predicting benign or pathogenic variants from the ClinVar database and correlation with deep mutational scanning assays – and at predicting a spectrum of pathogenicity across the proteome – by identifying patients with severe developmental disorders amongst controls based on de novo variants. A detailed analysis of severe developmental disorders cohorts was performed with the objective of identifying putative disease-causing variants or genes. Our strategy was two pronged. On the one hand, identify pathogenic variants based on a popEVE score threshold; this strategy can be applied to any patient without the need for a disease-cohort. And on the other hand, take advantage of the cohort structure and identify disease causing genes by performing a burden test of pathogenic variants at the gene level across the cohort. The results of both approaches were compared to state-of-art-approaches and putative variants were explored from a structural and functional perspective.

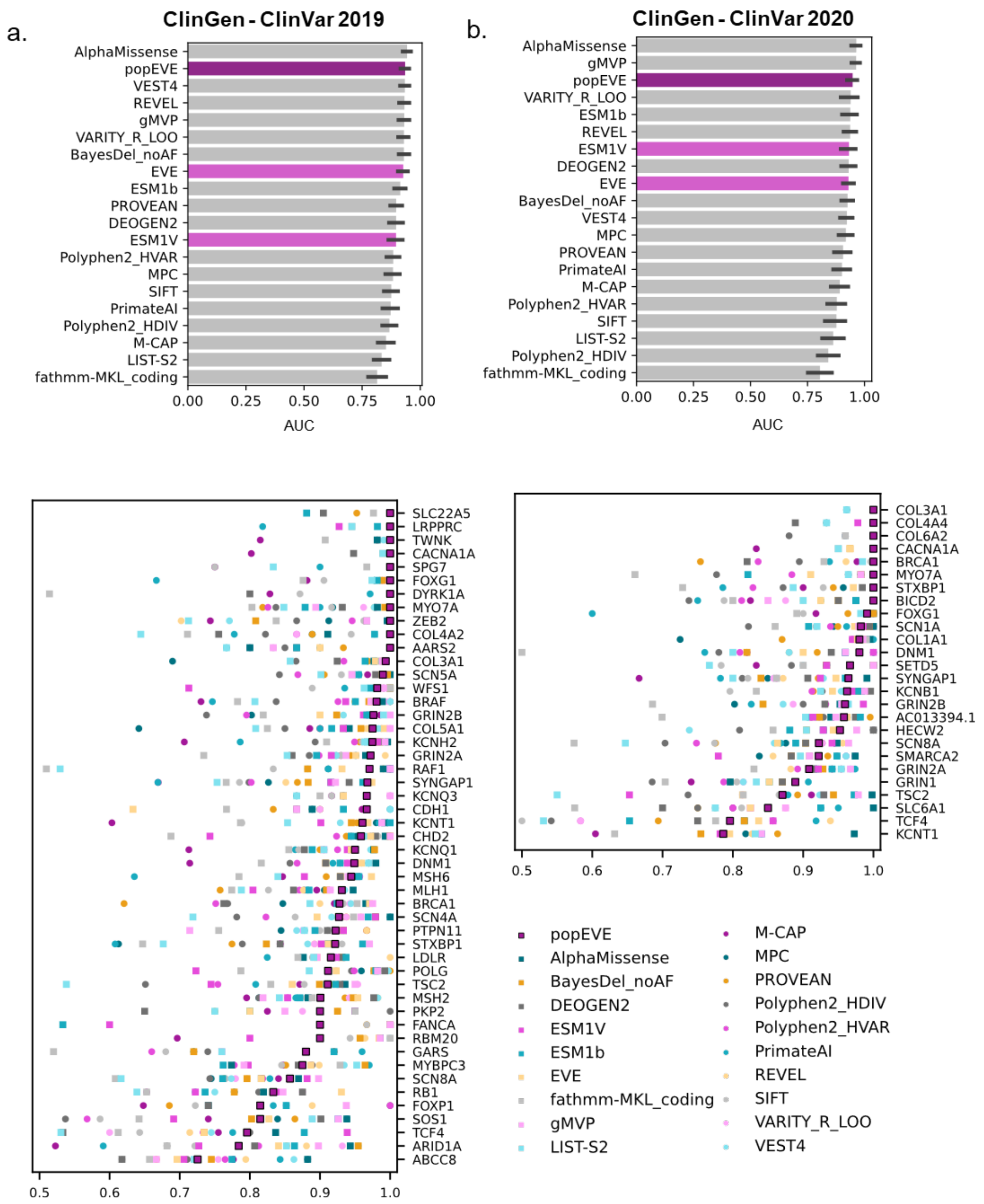


**Supplementary Figure 2. Performance summary for separating Benign/Likely Benign from Pathogenic/Likely Pathogenic ClinVar labels.** Assessing the performance of popEVE and popular supervised and unsupervised variant effect prediction models on individual genes that have at least 5 benign and 5 pathogenic variants from the ClinGen curation[1] of a. ClinVar 2019 and b. ClinVar 2020, using the area under the receiver-operating curve. The ClinGen dataset attempts to address data leakage in the estimation of performance of supervised methods by removing ClinVar variants used in training. This test lacks the resolution to distinguish state-of-the-art models. This is highlighted by the fact the ranking of AUCs in the ClinGen 2020 and ClinGen 2019 significantly changes.

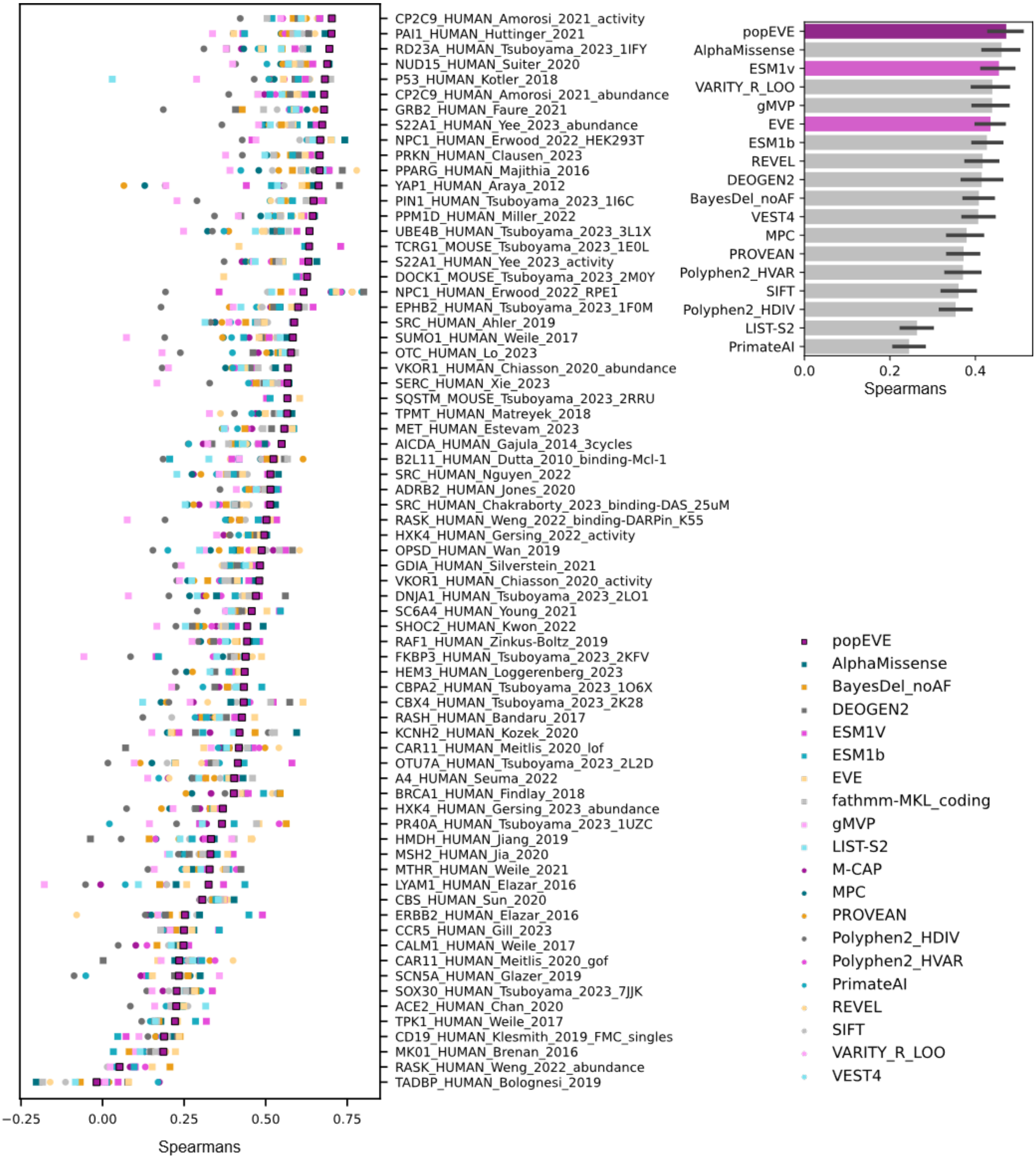


**Supplementary Figure 3. Correlation of computational variant effect predicting models with high-throughput experimental assays.** Assessing the performance of popEVE compared to popular supervised and unsupervised variant effect prediction models when compared to high-throughput functional assays on human genes (from ProteinGym[2]). On average popEVE outperforms other models.

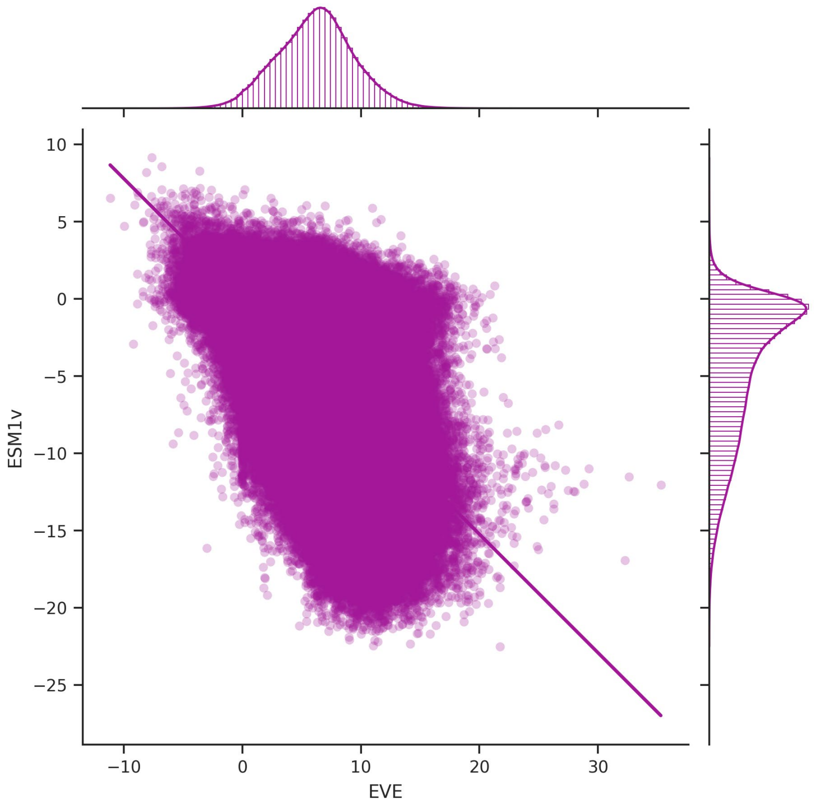


**Supplementary Figure 4. Correlation between EVE and Esm1v scores.** Scores for a random sample of 1 million variants across the human proteome. Pearson correlation is low – 0.55 (p-value=0.0).

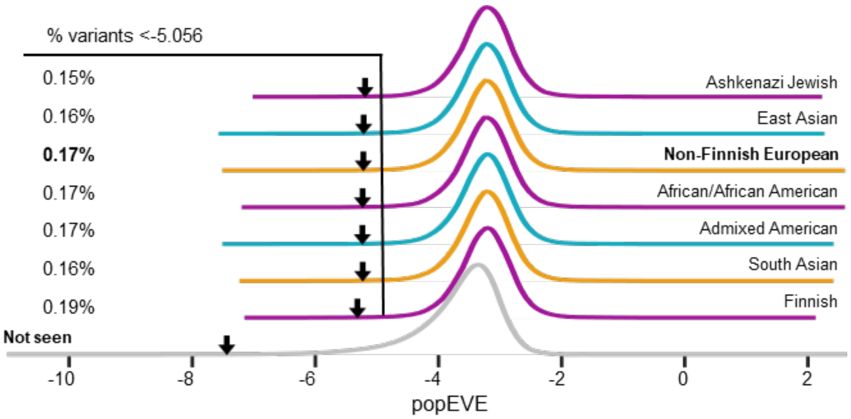


**Supplementary Figure 5. popEVE shows minimal population bias across diverse ancestries.** The distribution of popEVE scores for rare variants (AF<0.01) is consistent across populations found in gnomAD[4], indicating that despite using primarily non-Finnish European subjects for score adjustment there is no population bias. Variants not seen any gnomAD population are in grey. The 99.9% percentile for each distribution is marked with an arrow.

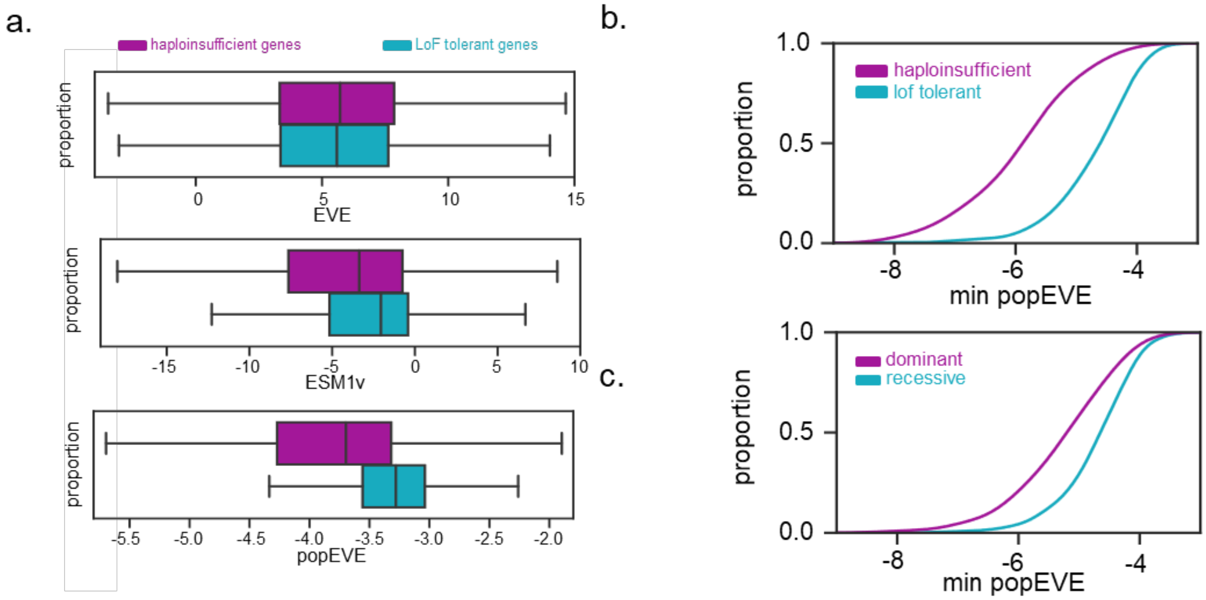


**Supplementary Figure 6. popEVE captures tail of pathogenic variants in genes involved in disease.** a. Model score distributions for single-edit distance substitutions in genes that are haploinsufficient[5] and LoF tolerant[6] (continuation of Fig. 1b). For EVE alone, the scores of these two sets of genes appear near identical (KS=0.04, pvalue=0). For ESM1v the deleterious end of the LoF intolerant gene scores is slightly denser, but both distributions have a substantial overlap between their inner quartiles. For popEVE, most variants in haploinsufficient genes[17] are more pathogenic than those in LoF tolerant genes[6]. b. Minimum, *i.e.* the most deleterious, popEVE score per gene can be used as a measure of gene constraint to distinguish between ClinGen haploinsufficient genes (n=186) and homozygous LoF tolerant genes (n=263) (KS=0.59, p=3e-40) [5,6]c. Minimum popEVE scores for genes with dominant (n=621) and recessive (n=1043) inheritance. Genes with dominant inheritance patterns have more pathogenic scores than genes with recessive inheritance (KS=0.32, p=1e-36)[7,8].

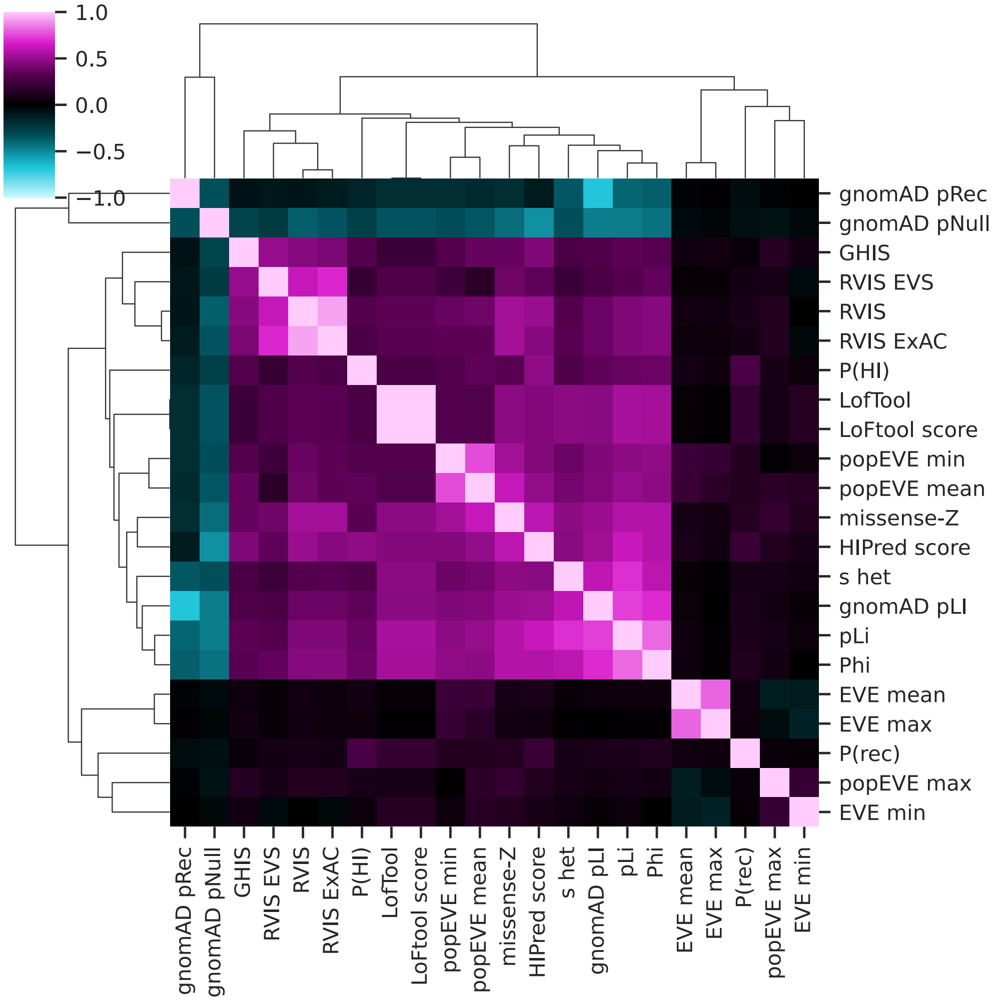


**Supplementary Figure 7. Correlation between popEVE gene-level statistics and gene-level constraint measures.** Pearson correlation between gene-level measures of constraint[3] and EVE and popEVE minimum, maximum and mean score per gene. We find poor correlation between popEVE and gene-level constraint measures, except for MissenseZ[4] and popEVE mean, with pearson = 0.61 (p-value = 0.0).

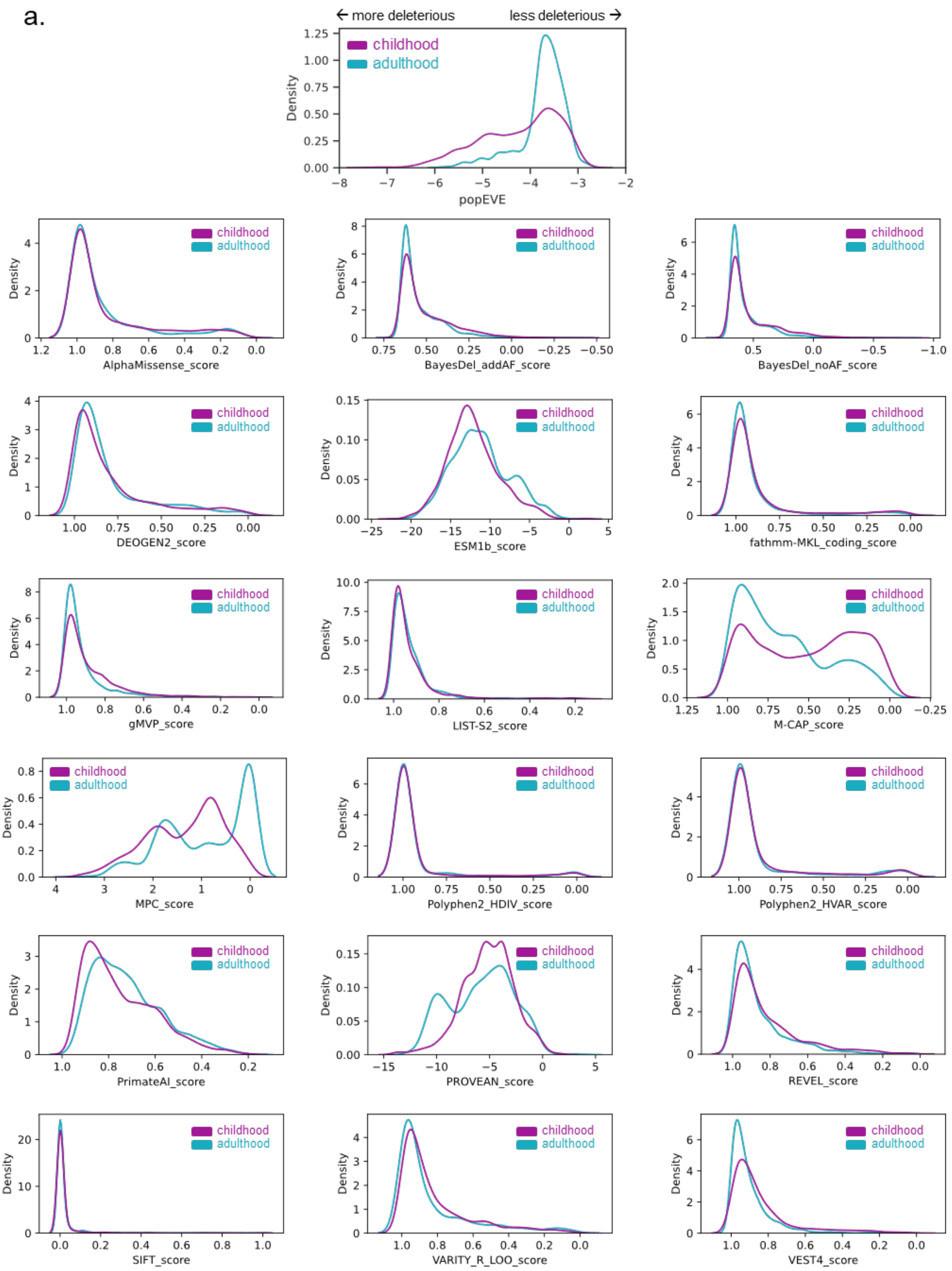
a.
← more deleterious
less deleterious →
childhood
adulthood
Density
popEVE
AlphaMissense_score
BayesDel_addAF_score
BayesDel_noAF_score
DEOGEN2_score
ESM1b_score
fathmm-MKL_coding_score
gMVP_score
LIST-S2_score
M-CAP_score
MPC_score
Polyphen2_HDIV_score
Polyphen2_HVAR_score
PrimateAI_score
PROVEAN_score
REVEL_score
SIFT_score
VARITY_R_LOO_score
VEST4_score

b.

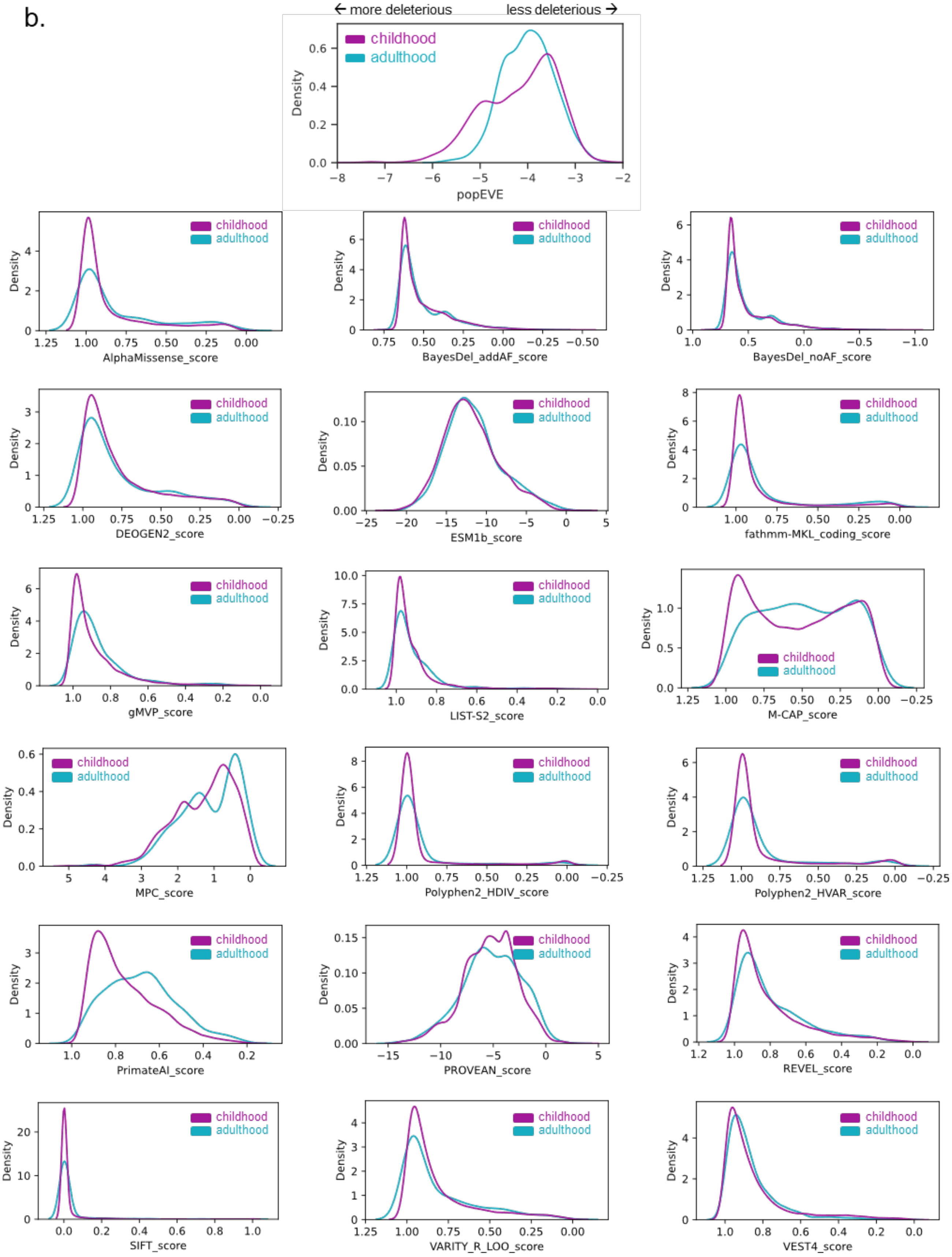

← more deleterious
less deleterious →
childhood
adulthood
Density
popEVE
AlphaMissense_score
BayesDel_addAF_score
BayesDel_noAF_score
DEOGEN2_score
ESM1b_score
fathmm-MKL_coding_score
gMVP_score
LIST-S2_score
M-CAP_score
MPC_score
Polyphen2_HDIV_score
Polyphen2_HVAR_score
PrimateAI_score
PROVEAN_score
REVEL_score
SIFT_score
VARITY_R_LOO_score
VEST4_score

**Supplementary Figure 8. Distributions of ClinVar pathogenic variants in genes associated with onset and death in childhood or adulthood.** Score distributions for various models of ClinVar pathogenic variants (with at least 1 star curation rating) in phenotypes associated with (a) death and (b) onset in childhood versus adulthood.

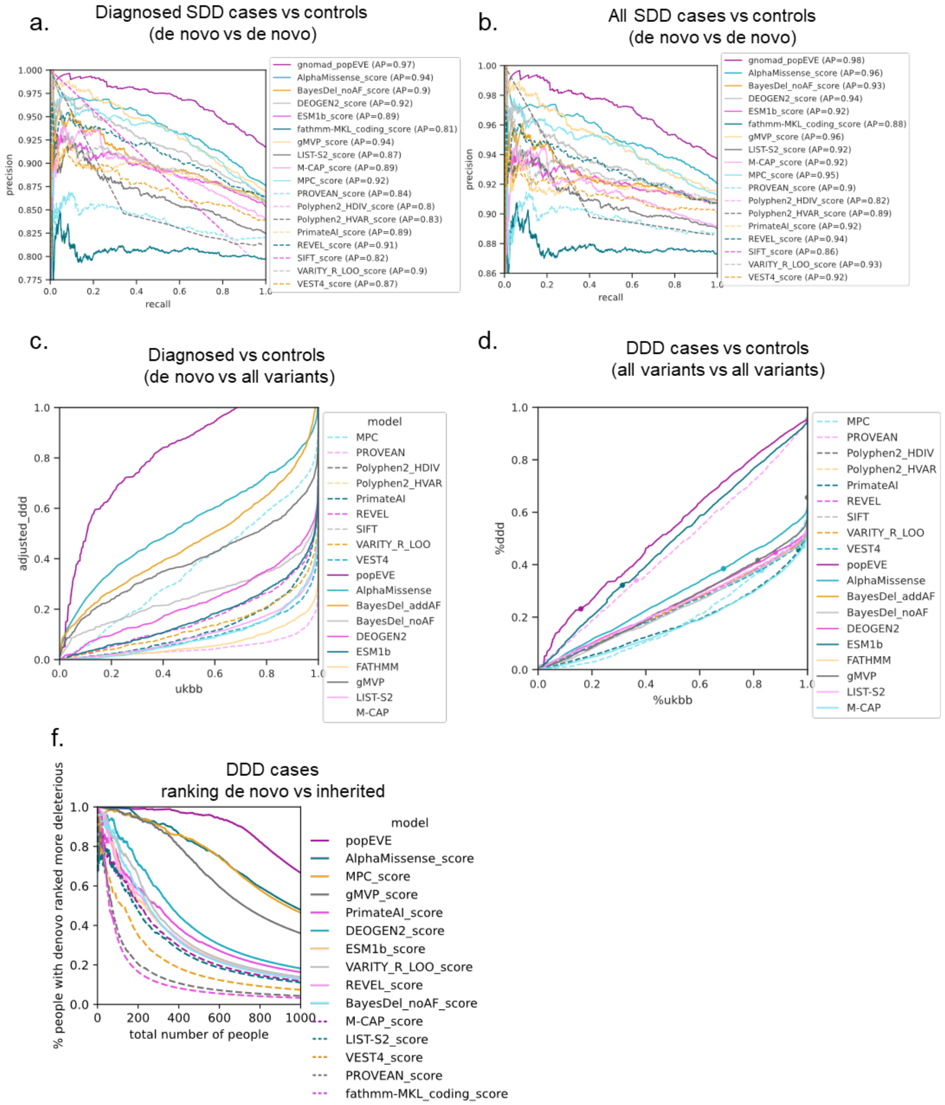


**Supplementary Figure 9. popEVE is better at separating developmental disorder cases from healthy controls than other state-of-the-art models.**

a. Extension of Figure 2c with all models - popEVE is better at separating "diagnosed" SDD cases whose disorder is likely to be caused by a de novo missense variant (cases with at least one missense variant in a known developmental disorder gene) from controls[8] than other state of art variant effect predictors with an average precision of 97%. b. Precision recall for "high-confidence diagnosed" SDD cases (at least one de novo missense in a gene discovered by DeNovoWEST in the same cohort). c. Precision recall for all cases vs controls. d. Extension of Figure 2d with all models - popEVE recalls more SDD cases (with at least one missense variant in DNW-discovered genes) without overpredicting pathogenicity in healthy controls from UKBB. While popEVE recalls 50% of these individuals for only 16% of the UKBB, the next best model, Alpha Missense[12], predicts 92% of UKBB has a variant as pathogenic as 50% of this SDD subset. e. Extension of Figure 3b with all models - For each score threshold, we plot the percent of individuals with a de novo missense variant ranked as more deleterious than rare inherited variants. In individual cases, popEVE is better at ranking de novo mutations as more deleterious than rare inherited variants (MAF<0.01) than other models. f. extension of Figure 3c with all models - popEVE recalls more DDD cases regardless of de novo or inherited labels without overpredicting pathogenicity in UKBB than other models.

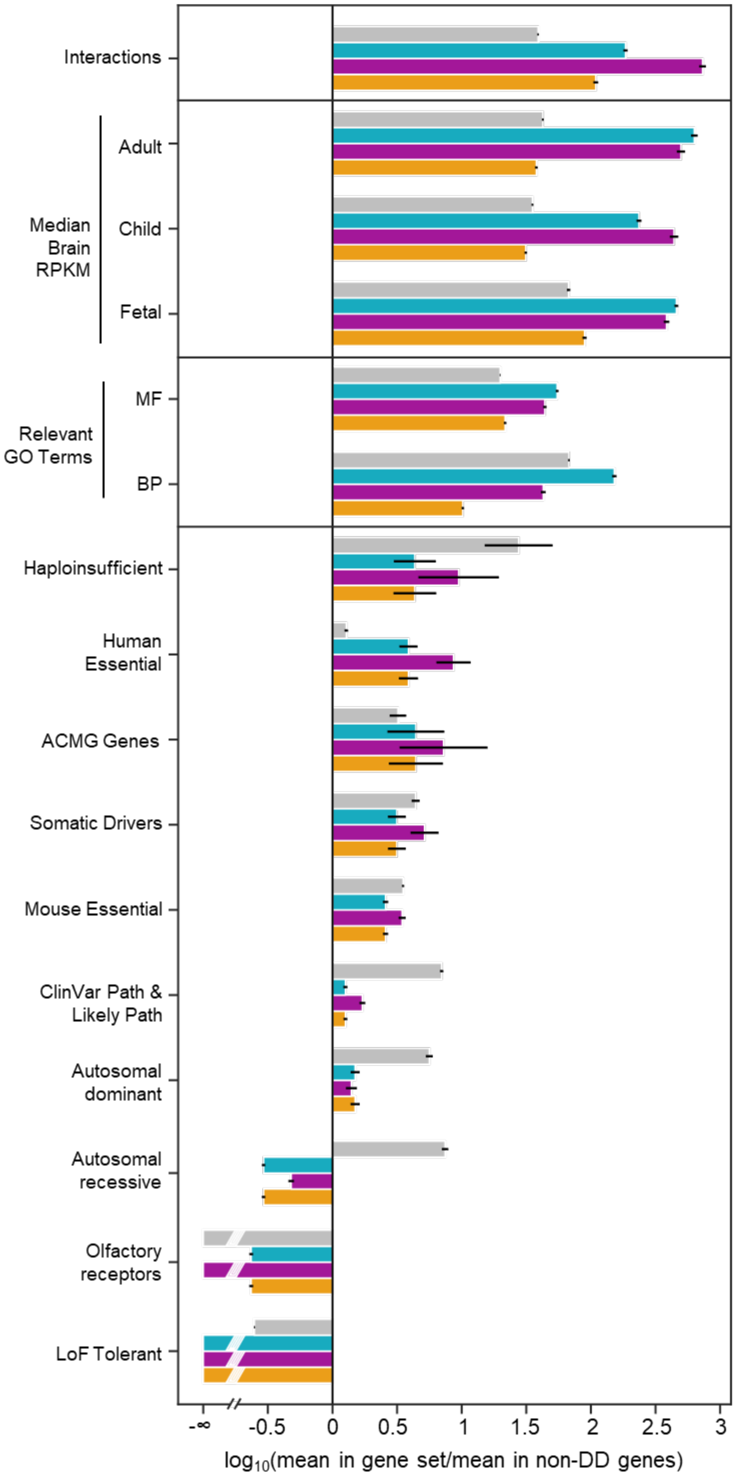


**Supplementary Figure 10. Functional enrichment of known and novel genes.**
Novel (from de novo SDD case variants and whole exome DDD variants) and known developmental disorder genes (from literature and previously discovered in the SDD cohort) show similar enrichment in properties known to differentiate known DD-genes from non-developmental disorder genes (95% CI from bootstrapping shown).

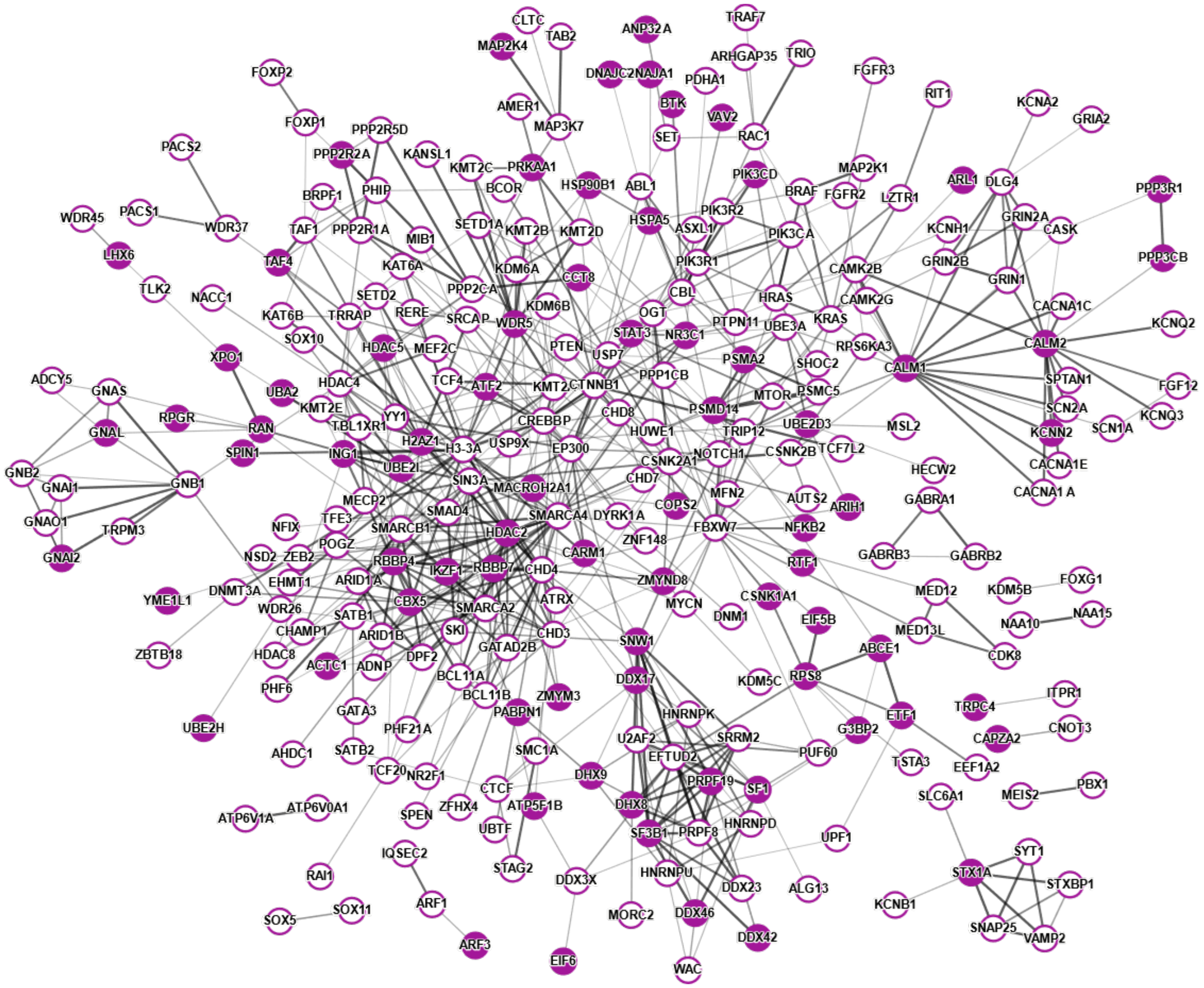


**Supplementary Figure 11. Functional network of previously known genes and newly discovered genes (extension of Fig. 3).**

Novel popEVE discovered genes are embedded into the network of previously-discovered disease associated genes from DDG2P and DeNovoWEST. Taking the set of 99.99 confidence threshold popEVE genes, we built a network using STRINGdb[13] ('experiments' and 'coexpression' at a medium 0.4 score threshold). Colored nodes are novel discoveries and white nodes are known disease-associated genes. These nodes were clustered into four clusters using k-means clustering.

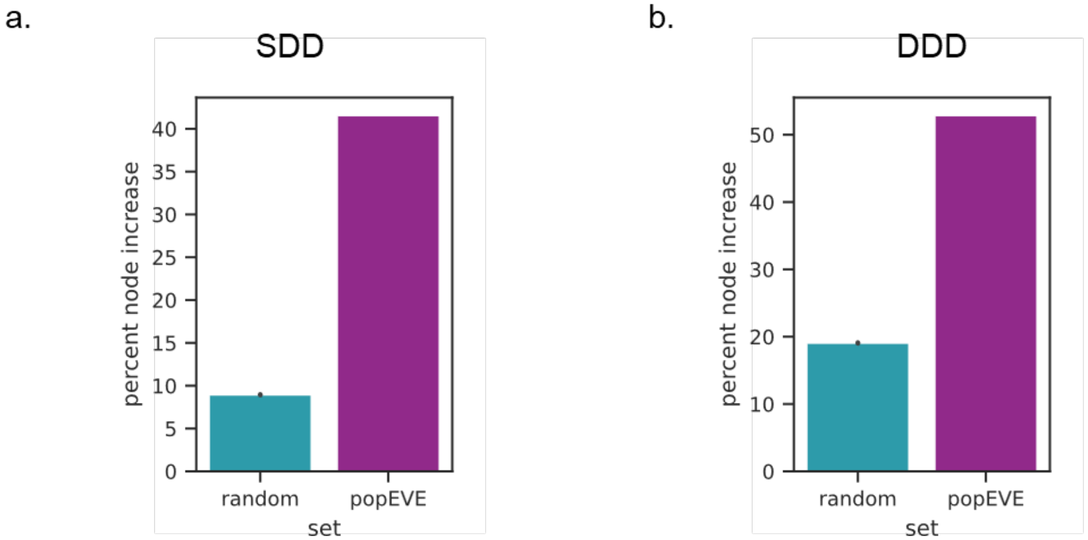


**Supplementary Figure 12. The DDG2P gene node connectivity increase of discoveries from the SDD and DDD compared to the null model.**
a. When added to a network of know developmental disorder genes, novel genes from the full SDD meta-cohort had a 42% increase in node degree as compared to random sets of the same number of genes which saw an average of 9% (with p=0, t-test). b. When added to a network of know developmental disorder genes, novel genes from the DDD sub-cohort had a 53% increase in node degree as compared to random sets of the same number of genes which saw an average of 19% (with p=0, t-test).
